# Designing an evidence-based Bayesian network for estimating the risk versus benefits of AstraZeneca COVID-19 vaccine

**DOI:** 10.1101/2021.10.28.21265588

**Authors:** Helen J. Mayfield, Colleen L. Lau, Jane E. Sinclair, Samuel J. Brown, Andrew Baird, John Litt, Aapeli Vuorinen, Kirsty R. Short, Michael Waller, Kerrie Mengersen

## Abstract

Uncertainty surrounding the risk of developing and dying from Thrombosis and Thromobocytopenia Syndrome (TTS) associated with the AstraZeneca (AZ) COVID-19 vaccine may contribute to vaccine hesitancy. A model is urgently needed to combine and effectively communicate the existing evidence on the risks versus benefits of the AZ vaccine. We developed a Bayesian network to consolidate the existing evidence on risks and benefits of the AZ vaccine, and parameterised the model using data from a range of empirical studies, government reports, and expert advisory groups. Expert judgement was used to interpret the available evidence and determine the structure of the model, relevant variables, data to be included, and how these data were used to inform the model.

The model can be used as a decision support tool to generate scenarios based on age, sex, virus variant and community transmission rates, making it a useful for individuals, clinicians, and researchers to assess the chances of different health outcomes. Model outputs include the risk of dying from TTS following the AZ COVID-19 vaccine, the risk of dying from COVID-19 or COVID-19-associated atypical severe blood clots under different scenarios. Although the model is focused on Australia, it can be easily adaptable to international settings by re-parameterising it with local data. This paper provides detailed description of the model-building methodology, which can used to expand the scope of the model to include other COVID-19 vaccines, booster doses, comorbidities and other health outcomes (e.g., long COVID) to ensure the model remains relevant in the face of constantly changing discussion on risks versus benefits of COVID-19 vaccination.

## 1 INTRODUCTION

Vaccine safety is a key consideration for public health managers, policy makers and the public. The controversy surrounding the safety of the AstraZeneca (AZ) COVID-19 vaccine (Vaxzevria) in relation to fatalities from rare, atypical severe blood clots (Thrombosis and Thrombocytopenia Syndrome [TTS]) (Greinacher et al. 2021) has contributed to vaccine hesitancy in many countries including Australia (Leask et al. 2021). This hesitancy is in-part due to the lack of access to comprehensive, up-to-date scientific information, displayed in a digestible and objective manner. The problem has been exacerbated by the continually-evolving information, with the publication of new scientific studies and agency reports. Moreover, many scientific studies often address only part of the overall puzzle. The lack of aggregated information presents challenges not only to members the public, but also to the clinicians who are tasked with helping patients make an informed decision about the AZ vaccine. Delving into the extensive and constantly-changing scientific literature is also beyond the resources of most public health practitioners and policy makers who need to make decisions in a highly dynamic environment. Similar challenges face epidemiologists and other scientists who need to assess and account for vaccine safety in related studies.

While there have been attempts to compile results into a meaningful comparison with respect to the risks versus benefits of the AZ vaccine, these have typically been confined to specific scenarios, for example when community transmission of SARS-CoV-2 in Australia was low (MacIntyre et al. 2021). A more flexible framework is therefore urgently needed to effectively combine and communicate the existing evidence surrounding the risks versus benefits of vaccines such as the AZ vaccine. It is crucial that this framework is both transparent in its assumptions and data sources, as well as easily updatable to account for new evidence and changes in the pandemic landscape, such as new variants, changes in vaccine effectiveness, or fluctuations in the rates of community transmission. The framework must also be able to incorporate data from a wide range of sources, and in different formats.

Bayesian network (BN) modelling (Fenton et al. 2013) is well suited for implementing such a framework. The transparency and flexibility of BNs for integrating different data sources (Marcot et al. 2001; Uusitalo 2007; Bertone et al. 2020) has seen them used for a variety of different analyses relating to COVID-19, such as examining the limitations of contact tracing (McLachlan et al. 2020), using expert-elicited data for interpretation of SARS-CoV-2 testing (Wu et al. 2021) and estimating SARS-CoV-2 infection and fatality rates (Neil et al. 2020). BNs can be designed as causal models and are easily interpretable, which makes them suitable for use in decision-support contexts, for example those developed by Fenton et al. to illustrate the need for more random testing of community members.

The COVID-19 Risk Calculator (CoRiCal) was developed to address the need for a user-friendly risk-benefit analysis tool to assist clinicians and the public to make informed decisions about COVID-19 vaccinations (Immunisation Coalition 2021; Lau et al. 2021). The specific objectives of the model were to estimate and compare: i) the risk of developing and dying from TTS following AZ vaccine; ii) the background risk of developing and dying from atypical severe blood clots (cerebral venous sinus thrombosis [CVST] and portal vein thrombosis [PVT]) in the general population; iii) the risk of developing and dying from COVID-19-related atypical severe blood clots (CVST or PVT); and iv) the risk of SARS-CoV-2 infection and related deaths under different transmission intensities (Lau et al. 2021). In this paper, we describe in detail the methods used to design and validate the BN model that integrates the best available evidence to compare the risks versus benefits of the AZ vaccine in the Australian population.

## 2 METHODS

### 2.1 Modelling Approach

The modelling process was a hybrid evidence-driven and expert-led approach (Figure 1). This process was modified from the approach used by Ticehurst et al. (2011), and was chosen as an efficient way of creating a useful and evidence-based model in as short a timeframe as possible. Six subject matter experts with experience in virology (KRS, JES), clinical practice (AB, CLL, JL), biostatistics (MW, KM), and infectious disease epidemiology (CLL, JL) were involved in gathering and interpreting available information. The modelling team (HJM, KM, CLL, JS) facilitated the design process and implemented the model using the GeNIe Bayesian network modelling software version 3.0.5703 (Bayes Fusion 2019).

**Figure 1.**
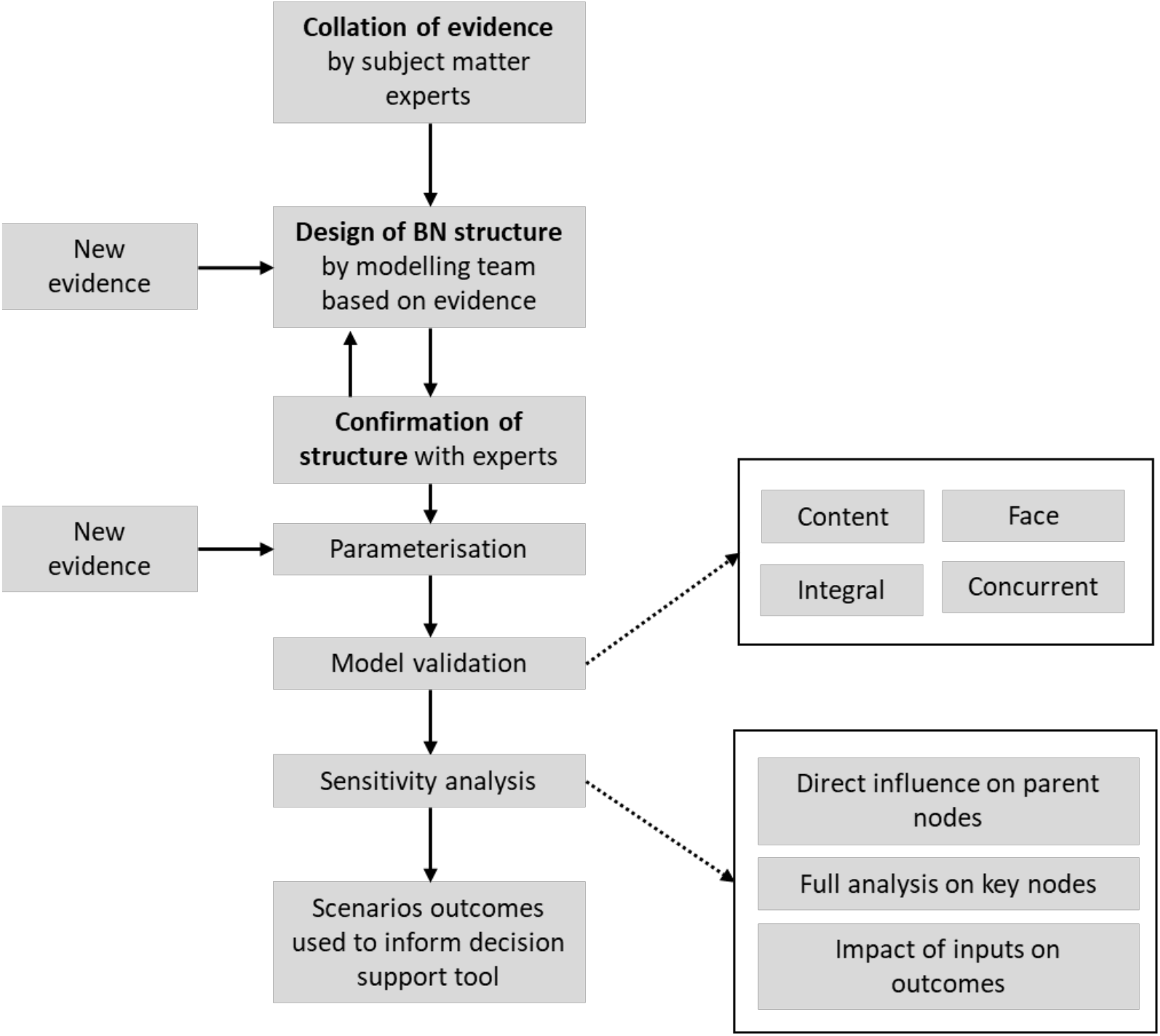
Model design process used for implementing the CoRiCal Bayesian network.

Evidence was initially collected for the incidence of the adverse events following immunisation (AEFI) for both the AZ and Pfizer vaccines, as well as for a range of adverse outcomes and complications from COVID-19 (including death), and comorbidities (e.g., obesity, diabetes) that could influence COVID-19-related outcomes. We selected CVST and PVT to provide a comparator to TTS which occurs after administration of a drug or vaccine. CVST and PVT are atypical severe blood clots that can occur at a background rate in the general population (Søgaard et al, 2016; Ageno et al., 2017; Kristoffersen et al., 2020) and have also been reported in COVID-19 patients (Taquet et al., 2021).

The scope of the model was narrowed in response to the changing pandemic landscape, where hesitancy for the AZ vaccine was growing, despite it being the only vaccine available to older age groups at the time of model development. To facilitate the urgent need for decision-support tools for clinicians and the general public, analysis of the Pfizer vaccine, comorbidities, and other COVID-19-related outcomes (e.g., ICU admission, long COVID) were excluded from this initial model. The results presented here therefore focus on the i) the risk of developing and dying from TTS following AZ vaccine; ii) the background risk of developing and dying from atypical severe blood clots iii) the risk of developing and dying from COVID-19-related atypical severe blood clots (CVST or PVT) and iv) the risk of SARS-CoV-2 infection and related deaths under different transmission intensities.

### 2.2 Data Sources

Three main sources of evidence were considered: published literature, publicly available government data/reports and professional expert advisory groups such as Thrombosis and Haematology Society of Australia and New Zealand (THANZ). In Australia, the Commonwealth Government is guided by an expert subcommittee of haematologists from THANZ and vaccine experts to adjudicate on whether cases of AEFI related to the AZ vaccine were classified as TTS. All publicly available data used in this study was sourced from official government websites (Section 3.1).

### 2.3 Bayesian Networks

BN models provide a visual and probabilistic approach to integrating and analysing data (Fenton et al. 2013). In a BN, the system of interest is depicted as a directed acyclic graph (DAG) in which variables are represented as nodes and parent-child associations between variables are represented as arrows connecting the respective nodes. In a discrete BN, each node is categorised into, or defined by, a set of states that define the classes (e.g., male/female) or ranges (e.g., 1-19, 20-30, 30+ years) of the corresponding variable. The BN is then quantified by assigning probabilities to these states, conditional on the states of the parent nodes. The probability table for a node without parents is quantified using a prior distribution. The BN structure therefore ascribes a set of conditional independencies on the joint distribution of the variables. This allows a rich model to be designed by encoding various model assumptions about the relationships of variables, while only needing to assign conditional probability tables (CPTs) on a few variables at a time, specifically between a child node and its immediate parent nodes. The simplification of a BN as a connected set of CPTs, in which each node is dependent only on immediate parent nodes is a result of the Markov properties of the underlying DAG, allows for very flexible model structure, fast computation and the ability to use different information sources to inform different components of the system (Uusitalo 2007).

The features of a BN are illustrated in the example shown in Figure 2, which demonstrates one option for modelling the probability of dying from vaccine-induced TSS based on AZ dose, variant (either no doses, first dose or second dose) and age group. The outcome node, *Die from vaccine-induced TTS* has one parent node; *Vaccine-induced TTS*. The *Vaccine-induced TTS* node is itself a child node of the *AZ dose* and *Age group* nodes. The CPT for the outcome node is shown, giving the probability for each state of this node conditional on the state of the parent node (*Vaccine-induced TTS*).

**Figure 2.**
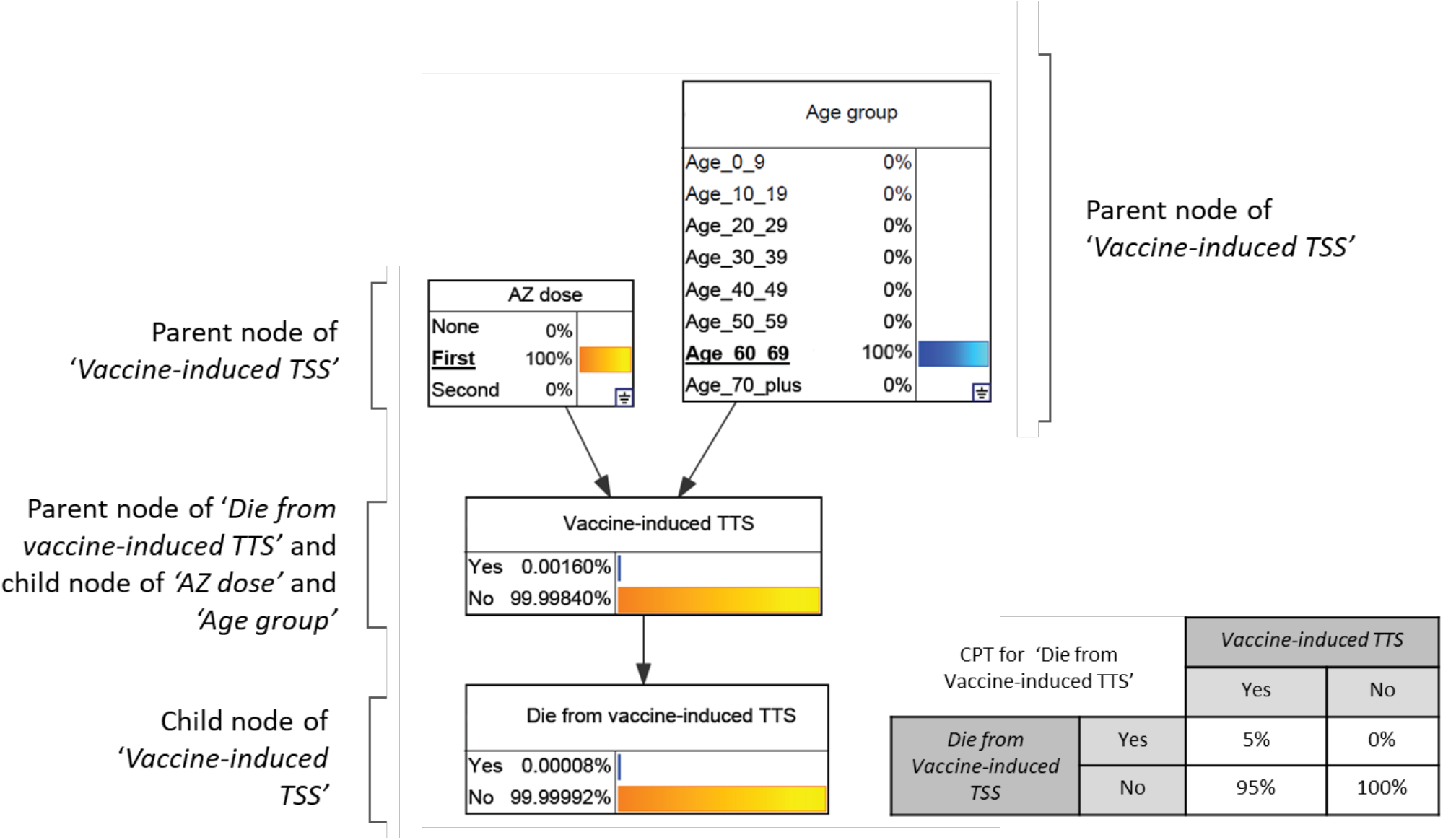
An example Bayesian network modelling the chance of dying from vaccine-induced TTS based on AZ dose and age group. Scenario shown is for the first dose of the AZ vaccine for a female aged between 60 and 69 old.

### 2.4 Model Design

#### 2.4.1 Model Structure

The structure of the BN model described here was developed in four stages. In the first stage, all authors agreed on the overall scope of the model. To accelerate development, it was decided to concurrently design and parameterise the model using three sub-models, based on subject matter. These were 1) risk of developing AZ vaccine-induced TTS, and background risk of developing and dying from atypical severe blood clots (CVST and PVT), i.e. in persons who have not received the AZ vaccine, nor been diagnosed with COVID-19; 2) risk of developing symptomatic COVID-19; and 3) risk of developing and dying from COVID-19 or COVID-19-related atypical severe blood clots (PVT and CVST). These three sub-models were then combined into a single model. The final conceptual model, which defined the structure of the resulting BN, represented the key relationships identified by the experts.

In the second stage of development, for each sub-model, the team of experts compiled a list of relevant variables, agreed on a list of reputable information sources for each variable, and defined the relationships between variables. Questionnaires were developed to formalise the evidence gathering (Supplementary S1 – Example questionnaire for evidence gathering) and prompt experts for relevant information (e.g. ‘Is there evidence to suggest that vaccine effectiveness differs by sex?’). After discussing the evidence with the experts, the answers from these questionnaires were used by the modelling team to design a draft conceptual model. This process was iterated over several weeks until agreement was reached between experts and modellers. The structure of the BN was modified as new evidence was identified or as updates became available. For example, when evidence emerged that the risk of COVID-19-related CVST and PVT differed by sex (Taquet et al. 2021), links between these nodes were added in the model.

In the third stage, the states of each node were defined. Most nodes in the model were binary, having only two possible states (yes/no; effective/ineffective), with age and community transmission being the only continuous variables requiring categorisation. Age was categorised into ten-year age brackets consistent with those used in the weekly Australian Technical Advisory Group on Immunisation (ATAGI) reports (ATAGI 2021a). Community transmission was categorised to be compatible with the ATAGI reported rates for low, medium and high transmission, as well as a baseline state (1000 cases per day) for three Australian states.

In the final stage, the model was critically evaluated to ensure that all variables and relationships could be informed by authoritative, quantitative evidence. Nodes and links in the BN were removed in the absence of evidence, or where the evidence suggested that the parent node had little effect on the outcome. For example, although the experts found evidence that males were at a higher risk of infection (NSW Government 2021), the difference between sexes was small and did not appear to have a substantive impact on the risk-benefit analysis; as highlighted above, this can be modified as further supporting information emerges. States of nodes were also examined in a similar manner and the states redefined where necessary according to the available evidence (for example, adding additional transmission rates).

#### 2.4.2 Parameterisation

Once the model structure was finalised, the available evidence was converted into a suitable format to quantify the CPTs. This involved defining the information in terms of conditional probabilities. For example, data reported as number of cases per 100,000 people, or infection rate over a certain time such as 16 weeks or six months, were converted into an equivalent probability and standardised to the same timeframes used in the model (6 weeks for background rates of CVST and PVT and 6 months for all COVID-19 related outcomes, including COVID-19-related atypical blood clots). CPT values were revised several times during the modelling process as new evidence or updated data became available.

Where evidence was available from more than one source, expert judgement was used to combine information or determine which source was most appropriate. For example, as there were limited Australian data on COVID-19-related CVST and PVT, this information was sourced from a large international study (Taquet et al. 2021). Expert judgement was also used if a particular source of evidence did not align with the model structure. For example, the age distribution of cases for the delta variant was sourced from daily reports by New South Wales (NSW) Health for the following age categories: 0-19, 5-year age groups from 20-69 years, and 70+. However, for the BN, the experts agreed that the 0-19 age category needed to be divided into 0-9 and 10-19. A 40%/60% split for the cases in the respective age groups was assumed, based on the age distribution of cases reported by the National Notifiable Disease Surveillance System (NNDSS) (NNDSS 2021).

### 2.5 Model Validation

Four key components of the BN model were inspected and critically evaluated by the modelling team: network structure, node discretisation, CPT parameterisation and overall model behaviour. The evaluation included the assessment of content, face, integral and concurrent validities (Pitchforth et al. 2013). Content and face validities were used to assess the model structure, the variables included and the relationships between them, and the discretisation of nodes within the network. All subject matter experts were provided with a step-by-step description of the final structure of the full model. They were then given the opportunity to discuss the assumptions and evaluate whether all relevant evidence from the literature had been included and appropriately represented in the model.

Integral validity was evaluated to confirm that the structure of the model reflected the design assumptions. Two statisticians (MW and KM) were provided with the assumptions and data used to calculate the CPTs. These values were then used to manually calculate the probabilities of various outcomes under different scenarios. These calculations were performed independently of model structure or parameterisation and the outcomes compared against model estimates for these same scenarios. Concurrent (or external) validity was used to assess the model behaviour. Scenarios were generated and the results were assessed for consistency with other common communication outlets such as ATAGI, for example the discretisation of age groups and definitions of low/medium/high transmission intensity (ATAGI 2021a). As described earlier, in some instances the choice was made to forego concurrent validity in favour of model parsimony based on the available evidence, for example excluding the reported weak relationship between sex and risk of infection as it would have little impact on the outcome but greatly increase the size of the CPT.

### 2.6 Model Sensitivity

Sensitivity analyses play a crucial role in assessing the robustness of the findings or conclusions of an analysis. They are an important way to assess the impact, effect or influence of key assumptions or variations—such as different methods of analysis, definitions of outcomes, protocol deviations, missing data, and outliers—on the overall conclusions of a model (Thabane et al. 2013). The sensitivity of the model was examined in three steps.

In the first step, for each intermediate and outcome node (i.e. all child nodes), the strength of influence of each parent node was calculated using the inbuilt function in the GeNie modeller software program (Bayes Fusion 2019). The strength of influence measures the difference in the probabilities of the target CPT based on changes in the probabilities of the parent CPT. A larger difference indicates a stronger influence. Two measures were considered, the average Euclidian difference and the maximum Euclidian difference (Supplementary S2 - Explanation of Euclidian Distance in BNs).

In the second step, this analysis was expanded to identify nodes that were most influential for the chance of developing symptomatic COVID-19 and the chance of dying from COVID-19. These two variables were selected as key nodes for analysis because both were highly connected (having three or more parents in the final model) and both had multiple indirect links with other nodes, so the route of influence is not easily determined by simply looking at the model structure. The most influential nodes were identified using the ‘Sensitivity Analysis’ function in the GeNie modeller software program (Bayes Fusion 2019).

In the third step, two separate analyses were carried out for all outcome nodes. The first examined which input nodes were most influential on the outcomes by adjusting the selected states for each node, for example selecting each age group one at a time, and then noting the estimated values of each outcome node. The second analysis for step three calculated and compared the minimum (worst-case) and maximum (best-case) estimates for each outcome node. The best-case and worst-case scenarios were identified by systematically adjusting the scenarios for each of the input nodes. The exception was that low, rather than no transmission was used for best-case scenarios for COVID-19-related outcomes as it was considered more meaningful. Under a zero-community transmission scenario, all outcome nodes other than *Die from background CVST (n16)* and *Die from background PVT (n17)* have a probability of zero, blocking any influence from the remaining input nodes.

## 3 RESULTS

### 3.1 Data Sources

The range of data sources used to design and parameterise the model are presented in **Table 1**. Evidence from Australian data was prioritised, however international studies were used to provide evidence on the rates of CVST and PVT, and vaccine effectiveness.

**Table 1.** Data sources used in designing and parameterising the model.

| Type of data | Variable | Data Source |
| --- | --- | --- |
| Government reports | Risk of developing and dying from vaccine-induced TTS | ATAGI weekly updates (ATAGI. 2021a) |
|  | Vaccine effectiveness against symptomatic infection and death | Australian Government report (delta variant)- Doherty Institute Modelling Report for National Cabinet, Table S2.5 (Doherty Institute, 2021)<br>UK Government report (alpha variant) - Vaccine effectiveness table, 16 July 2021. Public Health England (Vaccine Effectiveness Expert Panel, 2021) |
|  | Risk of developing symptomatic COVID-19 based on age group | Delta variant: (NSW Government 2021) - NSW COVID-19 cases data<br>Alpha Variant: (NNDSS 2021) National Notifiable Diseases Surveillance System public datasets |
| Peer-reviewed publications | Background chance of developing and dying from atypical severe blood clots (CVST or PVT) | CVST: (Kristoffersen et al. 2020) Incidence and Mortality of Cerebral Venous Thrombosis in a Norwegian Population<br>PVT:(Ageno et al. 2017) Incidence rates and case fatality rates of portal vein thrombosis and Budd-Chiari Syndrome; (Søgaard et al. 2016) Survival after splanchnic vein thrombosis: A 20-year nationwide cohort study. |
|  | Risk of developing and dying from COVID-19 related atypical severe blood clots (CVST and PVT) | (Taquet et al. 2021) Cerebral venous thrombosis and portal vein thrombosis: a retrospective cohort study of 537,913 COVID-19 cases. |
| Professional advisory group | Risk of developing and dying from TTS | ATAGI weekly updates (ATAGI. 2021a)<br>Thrombosis and Haematology Society of Australia and New Zealand (THANZ) Vaccine-induced Immune Thrombotic Thrombocytopenia (VITT) advisory group |
| Calculated | Risk of infection depending on age and variant | Calculated using formula, based on data from parent nodes |
|  | Risk of symptomatic COVID-19 infection under current transmission and vaccination status | Calculated using formula, based on data from parent nodes |

### 3.2 Model Description

The first sub-model, risks of developing and dying from background atypical severe blood clots (CVST and PVT), is shown in Figure 3. The risk of developing TTS associated with the AZ vaccine differs by age group and whether first or second dose of AZ is being considered (ATAGI 2021b). In this model, the risk of TTS after the first and second *AZ dose (n1)* were treated as independent events (risk of TTS after the first dose or after the second dose). For a population level analysis of total cases and deaths from TTS, a cumulative definition can be used where the risk of TTS after the second dose is the combined risk of both doses. *Die from vaccine-induced TTS (n15)* represents the proportion of TTS cases who die as a result (case fatality rate). The background rates of CVST and PVT were calculated over 6 weeks to be comparable to the timeframe in which TTS is likely to occur after the AZ vaccine.

**Figure 3.**
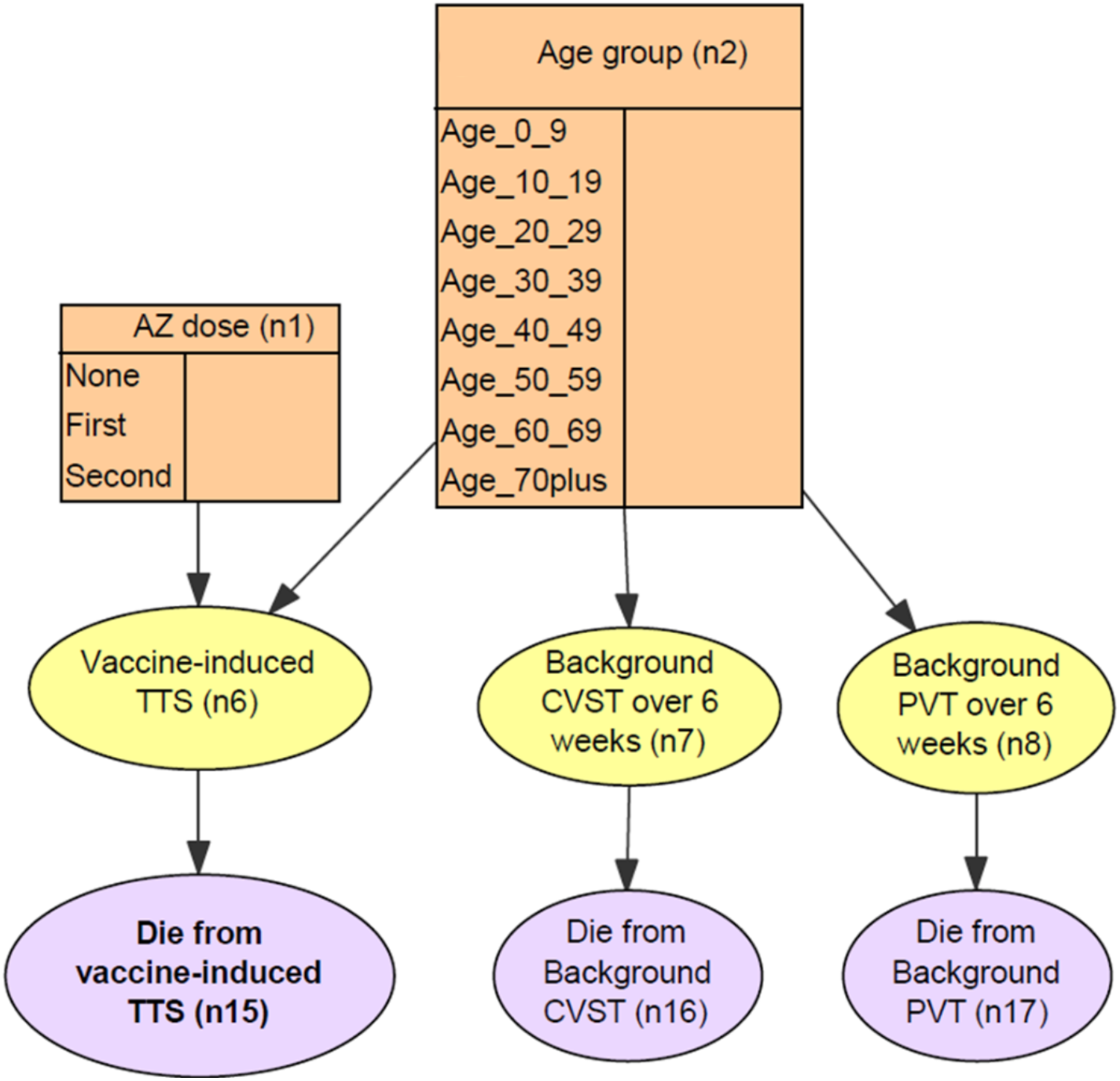
Conceptual model for sub-model 1: Risk of developing and dying from i) vaccine-associated TTS, and ii) background atypical severe blood clots (CVST and PVT), i.e. in those who have not received the AZ vaccine and have not been infected with SARS-CoV-2.

The second sub-model (Figure 4) focuses on the risk of developing symptomatic COVID-19 based on age group, SARS-CoV-2 variant and vaccine effectiveness. To enable direct comparison of risks of poor health outcomes versus benefits of the vaccine, all probabilities were calculated for a six-month period to reflect the estimated duration of protection from the AZ vaccine. Data were based on reported cases in different age groups from NSW (NSW Government 2021), which provided the best open source data available in Australia at the time of model development. In line with the dominant variant in Australia, data from June 2021 were used for the delta variant, and data prior to this date were used for the alpha/ancestral variant. Vaccine effectiveness was sourced separately for the delta variant (Doherty Institute 2021) and alpha/ancestral variant (Vaccine Effectiveness Expert Panel 2021).

**Figure 4.**
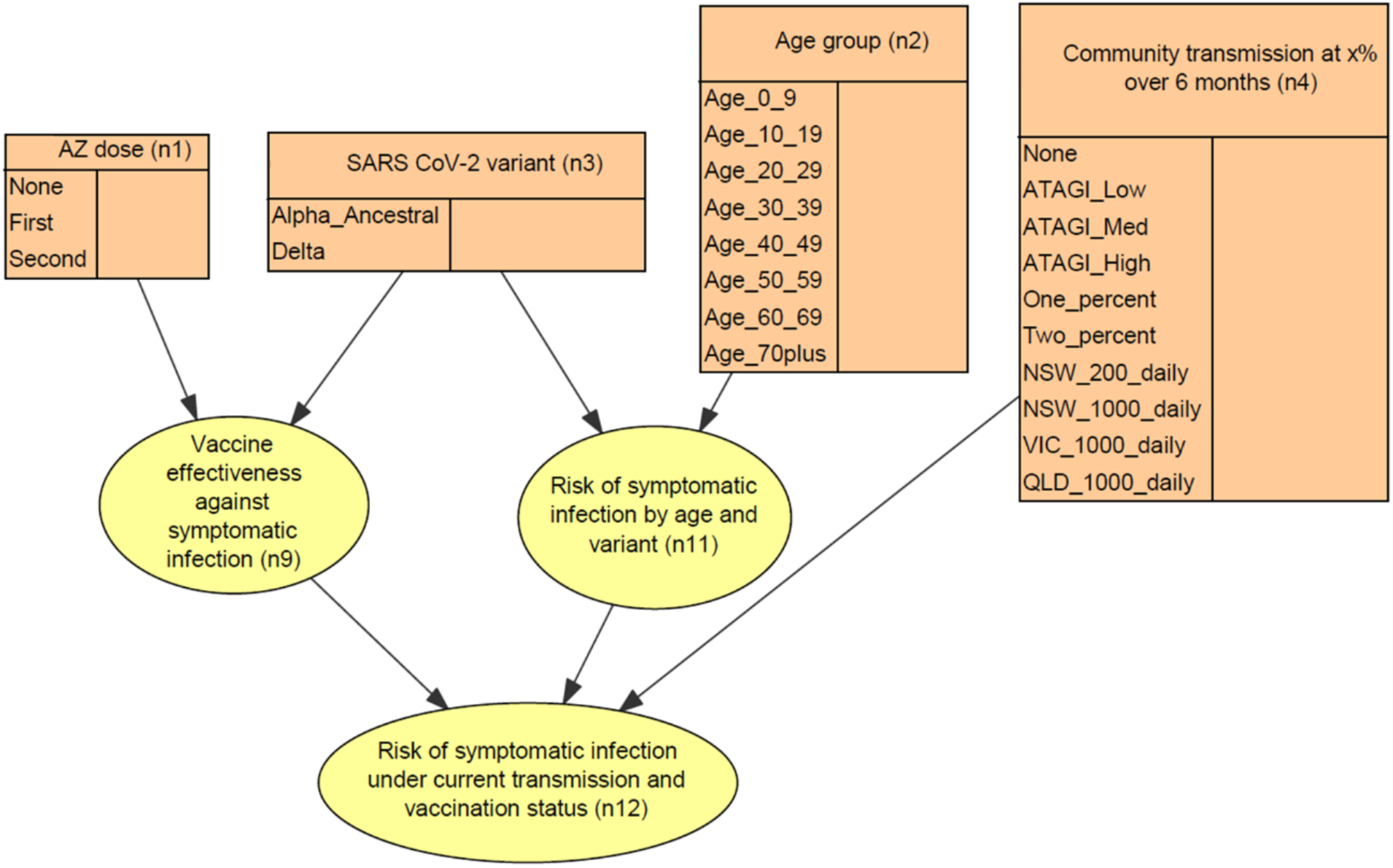
Conceptual model for sub-model 2: Risk of developing symptomatic COVID-19 depending on number of AstraZeneca vaccine doses received, SARS-CoV-2 variant, vaccine effectiness, age group, and level of community transmission.

To simplify the model structure and enable easier updates in the future, the relative risk of symptomatic infection by age and variant was calculated using an intermediate node (n11). The *Risk of symptomatic infection under current transmission and vaccination status (n12)* over a 6-month period was thus calculated for different transmission intensities that reflect various realistic scenarios. The intermediate step of calculating the *Risk of symptomatic infection by age and variant (n11)* was not necessary from a mathematical perspective, but was included to simplify the conceptual model and resulting CPTs. This reduces the number of different combinations in the resulting CPT and makes it easier for experts to quantify them with unique probabilities.

The final sub-model (Figure 5) collated evidence for the risk of dying from COVID-19, or from COVID-19-related CVST or PVT.

**Figure 5.**
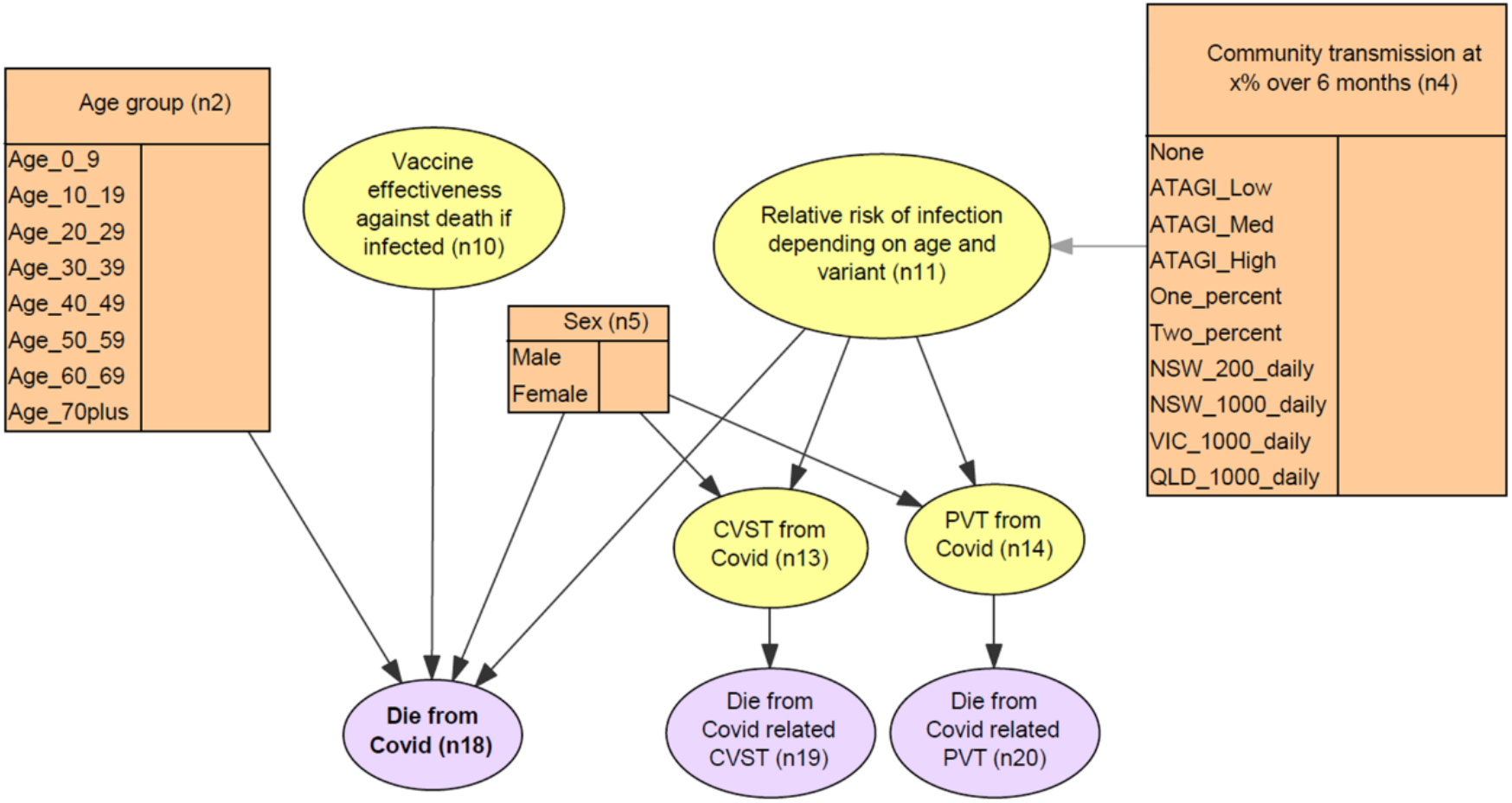
Conceptual model for sub-model 3: Dying from COVID-19 or COVID-19-related atypical severe blood clots, depending on age, sex, vaccine effectiveness, variant, and level of community transmission.

The final BN (Figure **6**) combines the three sub-models and integrates the current available evidence regarding the probability of *Dying from vaccine-induced TTS (n15), Dying from background CVST (n16)* or *Dying from background PVT (n17)*, overall probability of *Dying from COVID-19 (n18)*, and probability of *Dying from Covid-related CVST (n19)* or *Dying from Covid-related PVT (n20)*. Five input nodes – *AZ dose (n1), Age group (n2), SARS-CoV-2 variant (n3), Community transmission at x% over 6 months (n4)* and *Sex (n5)* were used to define the population and transmission scenarios.

**Figure 6.**
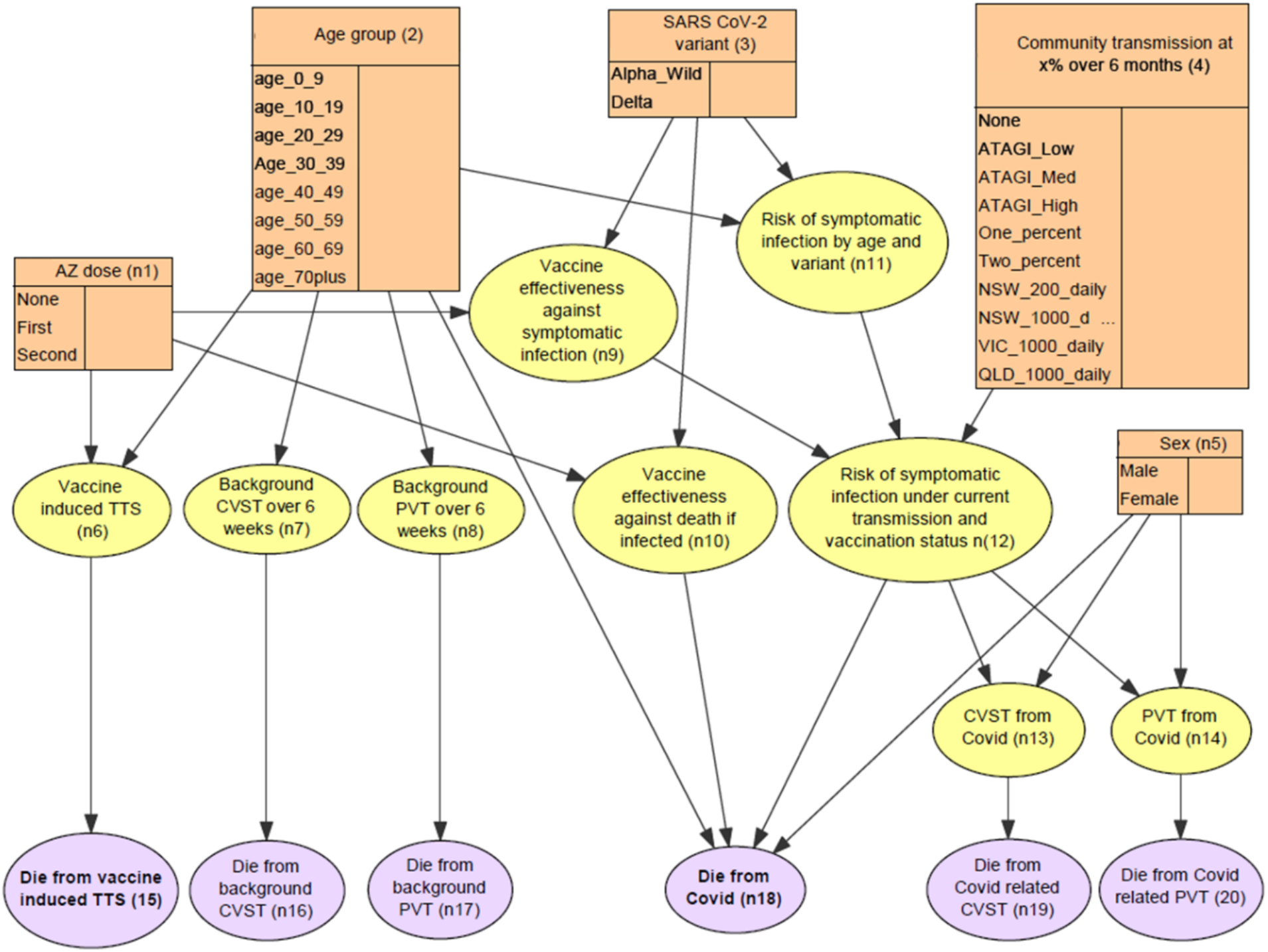
Bayesian network structure showing relationships between the income, outcome and intermediate nodes.

Several latent nodes were included as intermediate steps when calculating the probability of *Dying from COVID-19 (n18). Vaccine effectiveness against symptomatic infection (n9)* and *Vaccine effectiveness against death if infected (n10)* were modelled based on the *SARS-CoV-2 variant (n3)* (either alpha/ancestral or delta) and the *AZ dose (n1)* received (none, first or second)). At the time of this study, there was insufficient evidence to include the effects of the different vaccine schedules (e.g., shorter intervals between first and second doses) on vaccine effectiveness.

The *Risk of symptomatic infection by age and variant (n11)* was calculated assuming a community transmission rate of 10% over six months. This was then used to estimate the *Risk of symptomatic infection under current transmission and vaccination status (n12)* based on the *Community transmission at x% over 6 months (n4)* and *Vaccine effectiveness against symptomatic infection (n9)*. The final variables used to calculate the risk of *Dying from COVID-19 (n18)* were *Age (n2), Sex (n5)*, the *Risk of symptomatic infection under current transmission and vaccination status (n12)* and *Vaccine effectiveness against death if infected (n10)*. The remaining two outcome nodes were *Die from COVID-19-related CVST* (n19) and *Die from COVID-19-related PVT (n20)*, each dependent on developing *CVST from COVID-19 (n13)* or *PVT from COVID-19 (n14)*, respectively.

### 3.3 Parameterisation

Full probability tables for all nodes are given in Supplementary S3 – Values for conditional probability tables. Of the five input nodes, priors for two of these (sex and age group) were set to reflect the approximate distribution of the Australian population. For *Sex (n5)* a uniform distribution (50% males and 50% females) was adopted. The distribution for the *Age groups (n2)* was based on data from the Australian Bureau of Statistics (Australian Bureau of Statistics 2021). To reflect the COVID-19 situation in Australia at the time the analysis was carried out (August 2021), the priors for the *SARS-CoV-2 variant (n3)* were set as 95% delta variant and 5% alpha/ancestral variant. *AZ dose (n1)* was set to 30% unvaccinated, 35% received first dose only, and 35% received second dose. Priors for AZ dose can be updated as vaccine coverage increases. Uniform priors were used for the *Community transmission at x% over six months (n4)* - where x is the percent transmission at each state, as it is expected that a value will be selected for this state prior to running scenarios.

The default probabilities for the model are shown in Figure 7. Scenarios can be generated by selecting a single state for each input node (shown in orange). This evidence then propagates through the network to produce an estimate from each outcome. Additional scenarios can be generated to evaluate the effect of the vaccine once a person becomes infected with SARS-CoV-2 by setting node n12 to ‘Yes’. Probability values for a medium transmission scenario (n4) for the delta variant (n3) and a fully vaccinated population (n1) are given in Supplementary S4 – Probabilities for scenario -, as an example of scenario analysis.

**Figure 7.**
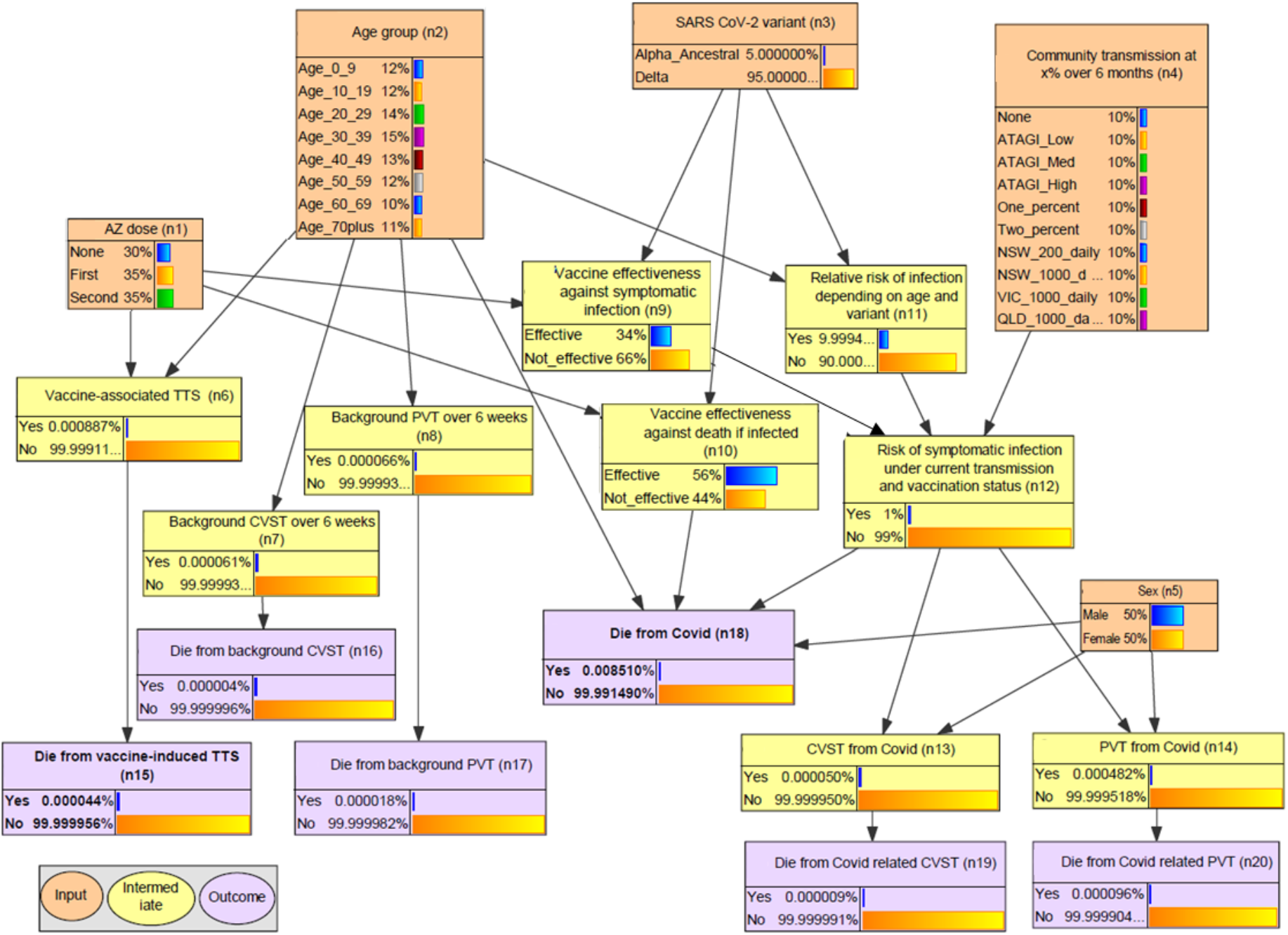
Parameterisation for the delta variant under a medium transmission scenario.

Of the 15 intermediate and outcome nodes, three (*Vaccine effectiveness against symptomatic infection [n9], Vaccine effectiveness against death if infected [n10]*, and relative *Risk of symptomatic infection by age and variant [n11]*) were calculated from data provided in government reports (Doherty Institute 2021; NSW Government 2021; Vaccine Effectiveness Expert Panel 2021). The case fatality rate for vaccine-induced TTS (n15) was sourced from ATAGI reports (ATAGI 2021b).

For the *Risk of symptomatic infection under current transmission and vaccination status (n12)*, the CPTs for any scenario in which *Vaccine effectiveness against symptomatic infection (n9)* equalled ‘Yes’ (i.e. the vaccine is effective) or relative *Risk of symptomatic infection by age and variant (n11)* equalled ‘No’ (i.e. there was no risk of infection) were parameterised at 100% probability of no infection (i.e. risk equals zero). For the remaining scenarios (i.e., the *Vaccine effectiveness against symptomatic infection (n9)* was ‘No’ and relative *Risk of symptomatic infection by age and variant (n11)* was ‘Yes’), the probability for each state of *Community transmission at x% over six months (n4)* was calculated as a proportion of the 10% baseline in node n11. For example, in node n12 the state ‘Two-percent’ represents a 2% community transmission rate over six months, which is one fifth of the 10% baseline assumed for node n11. Under this scenario, the value for the ‘Yes’ state in the CPT is therefore 0.2, or 20% of the assumed baseline. The remaining nine CPTS were populated directly with data from published literature, and modified for external consistency if necessary. A full explanation of all assumptions used in the calculations is given in Supplementary S5 – Assumptions.

### 3.4 Validation

All subject matter experts were satisfied that the final conceptual model structure sufficiently represented all relevant variables and relationships within the scope of the model, and that chosen states for each node were consistent with what could be parameterised based on the available evidence. Calculations made independently of the model predictions agreed with model predictions (Supplementary S6 - Manual calculations for validation), confirming the integral validity of the model structure in meeting the design assumptions. The results of the scenario testing (Supplementary S7 - Influence of inputs nodes on outcomes), which were also used for sensitivity analysis, confirmed that the model behaved as expected. For example, increasing the rate of community transmission increased the fatalities from COVID-19 and COVID-19-related atypical severe blood clots. Similarly, increasing the proportion of the population who were vaccinated decreased these same outcomes.

One potential anomaly revealed in this analysis was that for lower age groups, model estimates for dying from COVID-19-related atypical severe blood clots were higher than estimates of the overall probability of dying from COVID-19. Investigation revealed that this was a result of the data being taken from different sources, with the probability of blood clots being derived from UK data (Taquet et al. 2021) and the COVID-19 case fatality data from Australia (NSW Government 2021), where there had been relatively few COVID-19 fatalities at the time of writing. While this should not be considered as an error in the model, it has the potential to cause confusion when end-users are interpreting scenario results, which may reduce trust in the model estimates. Clear communication around the limitations and assumptions of the model are critical in helping to prevent these misunderstandings.

### 3.5 Sensitivity Analysis

The results of the strength-of-influence analysis comparing the relative influence of direct parent nodes on child nodes are shown in Table 2. Relative influence measures are denoted as average and maximum Euclidean distance (ED), where a larger ED indicates greater influence. For example, the analysis shows that a person’s age (average ED 0.013) was more influential than their sex (average ED 0.001) in determining their chance of *Dying from COVID-19 (n18)*. Age (ED 0.043) was also more influential than variant (ED 0.031) in determining the *Risk of symptomatic infection by age and variant (n11)*.

**Table 2.** Strength of influence of parent nodes on child nodes based on Euclidian distances (ED). Outcome nodes are shown in bold.

| Child | Parents | Average ED | Maximum ED |
| --- | --- | --- | --- |
| *Vaccine-induced TTS (n6)* | *Age group (n2)* | 1.00E-06 | 1.10E-05 |
|  | *AZ dose (n1)* | 1.60E-05 | 2.70E-05 |
| *Background CVST over 6 weeks (n7)* | *Age group (n2)* | 2.00E-07 | 4.00E-07 |
| *Background PVT over 6 weeks (n8)* | *Age group (n2)* | 1.00E-06 | 2.00E-06 |
| *Vaccine effectiveness against symptomatic infection (n9)* | *AZ dose (n1)* | 0.470 | 0.800 |
|  | *SARS-CoV-2 variant (n3)* | 0.153 | 0.270 |
| *Vaccine effectiveness against death if infected (n10)* | *AZ dose (n1)* | 0.617 | 0.950 |
|  | *SARS-CoV-2 variant (n3)* | 0.053 | 0.110 |
| *Risk of symptomatic infection by age and variant (n11)* | *Age group (n2)* | 0.044 | 0.117 |
|  | *SARS-CoV-2 variant (n3)* | 0.031 | 0.072 |
| *Risk of symptomatic infection under current transmission and vaccination status (n12)* | *Community transmission at x% over 6 months (n4)* | 0.052 | 0.576 |
|  | *Risk of symptomatic infection by age and variant (n11)* | 0.091 | 0.576 |
|  | *Vaccine effectiveness against symptomatic infection (n9)* | 0.091 | 0.576 |
| *CVST from Covid (n13)* | *Risk of symptomatic infection under current transmission and vaccination status (n12)* | 4.20E-05 | 5.40E-05 |
|  | *Sex (n5)* | 1.30E-05 | 2.50E-05 |
| *PVT from Covid (n14)* | *Risk of symptomatic infection under current transmission and vaccination status (n12)* | 4.01E-04 | 4.83E-04 |
|  | *Sex (n5)* | 8.20E-05 | 1.64E-04 |
| ***Die from vaccine induced TTS (n15)*** | *TTS (n6)* | 0.050 | 0.050 |
| ***Die from background CVST (n16)*** | *Background CVST over 6 weeks (n7)* | 0.070 | 0.070 |
| ***Die from background PVT (n17)*** | *Background PVT over 6 weeks (n8)* | 0.270 | 0.270 |
| ***Die from Covid (n18)*** | *Age group (n2)* | 0.013 | 0.217 |
|  | *Risk of symptomatic infection under current transmission and vaccination status (n12)* | 0.014 | 0.217 |
|  | *Sex (n5)* | 0.001 | 0.026 |
|  | *Vaccine effectiveness against death if infected (n10)* | 0.014 | 0.217 |
| ***Die from Covid related CVST (n19)*** | *CVST from Covid (n13)* | 0.174 | 0.174 |
| ***Die from Covid related PVT (n20)*** | *PVT from Covid (n14)* | 0.199 | 0.199 |

Figure 8 shows the results of the sensitivity analysis for the *Risk of symptomatic infection under current transmission and vaccination status (n12)*. The strength of influence is shown by the shading, with darker red shading representing a stronger influence. Nodes shown in grey have no influence on the target node (n12) because of the network structure. As expected, when all nodes were considered (rather than just the direct parents), *Community transmission at x% over 6 months (n4)* was highly influential on the *Risk of symptomatic infection under current transmission and vaccination status (n12)* (Figure 8). Of the input nodes, both *Age group (n2)* and the *AZ dose (n1)* had a strong influence on the *Risk of symptomatic infection under current transmission and vaccination status (n12)*.

**Figure 8.**
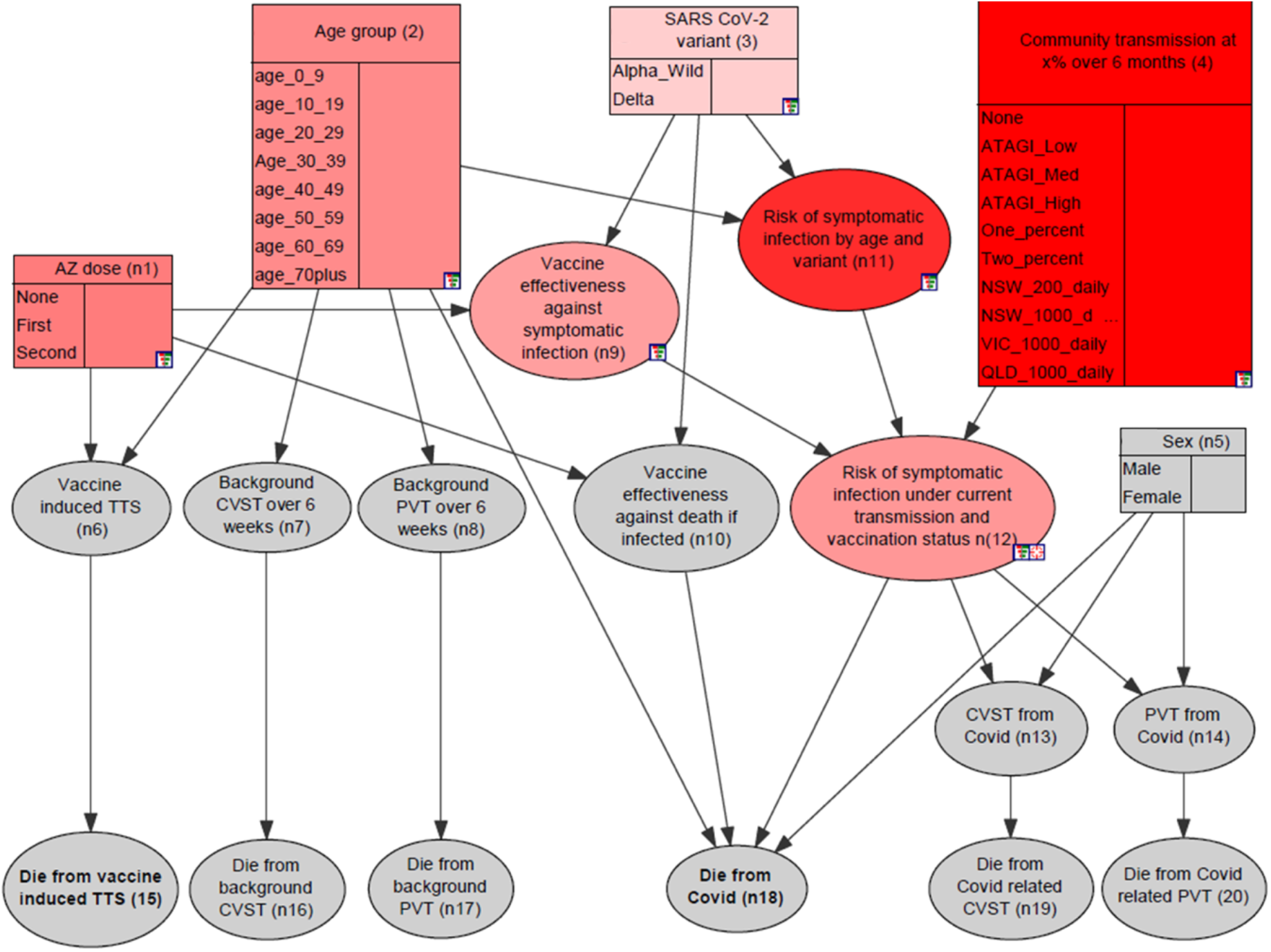
Sensitivity to findings for ‘Risk of symptomatic infection under current transmission and vaccination status (n12)’. Nodes with darker red shading have more influence than lighter shaded nodes. Grey nodes are not connected to the target node (n12) and therefore have no influence.

The equivalent analysis for *Die from Covid* (n18) in Figure 9, shows that *Age group (n2)* has a large influence. The *AZ dose (n1)* is shown as influential in both preventing *Risk of symptomatic infection under current transmission and vaccination status (n12)* and preventing *Dying from Covid (n18)* once infected. The stronger influence of *Age group (n2)* compared with *AZ dose (n1)* on the *Die from Covid (n18)* node is due to both the input data and the model structure, in that nodes directly linked to the target node will be more influential than variables linked indirectly.

**Figure 9.**
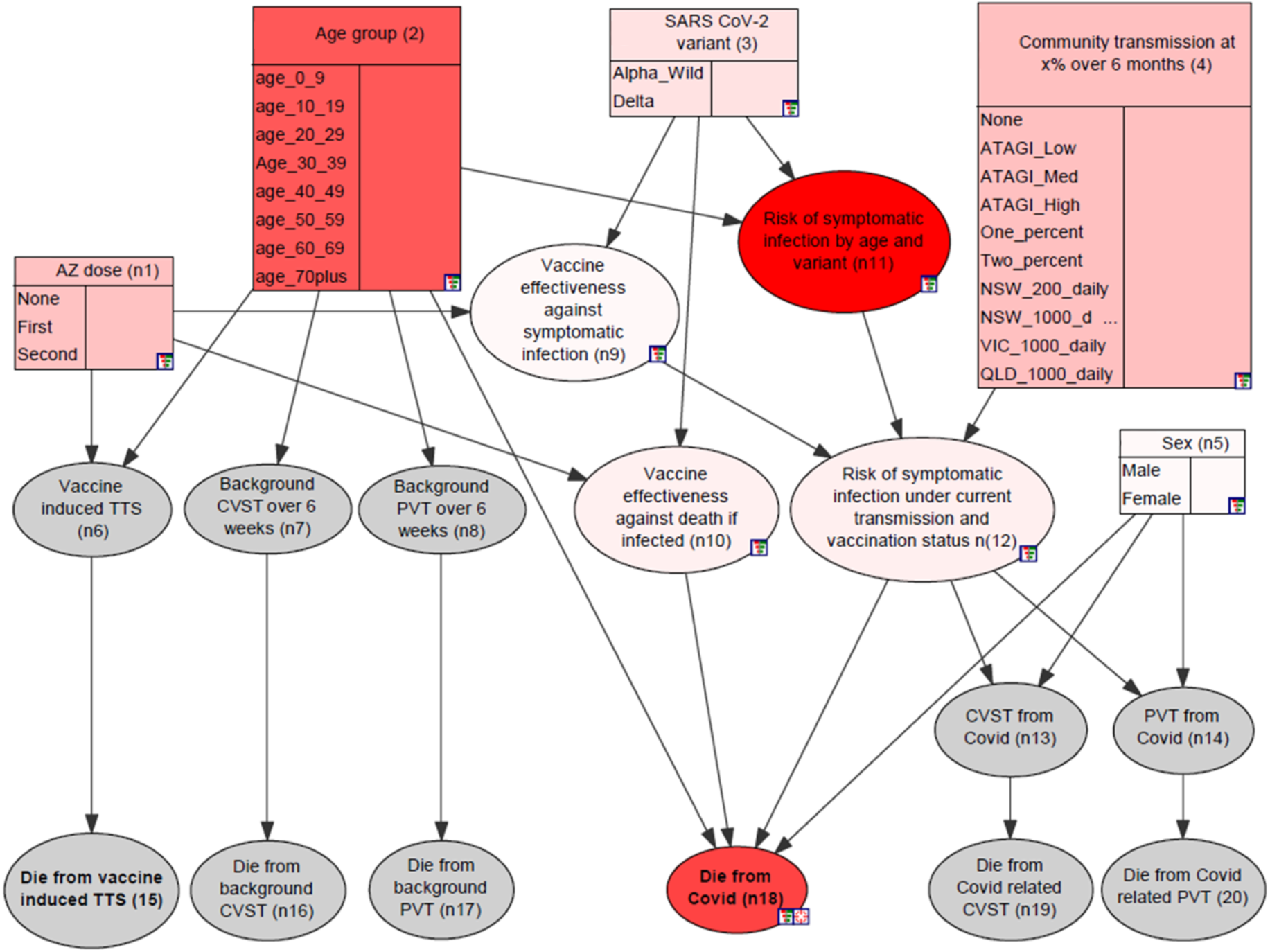
Sensitivity to findings for ‘Die from Covid (n18)’. Nodes with darker red shading have more influence than lighter shaded nodes. Grey nodes have no influence.

The results of alternating the states of individual input variables are provided in Table 3. The minimum and maximum values in Table 3 represent the largest and smallest feasible probabilities for each outcome node being ‘Yes’, when the respective input node is used to define a scenario. For example, when no evidence was set for any other input nodes, changing the values of *AZ dose (n1)* results in a maximum estimated probability of *Dying from COVID-19 (n18)* equivalent to 224 per million, and can be reduced to at most 8 deaths per million. *AZ dose (n1)* in these tables, and in Figures 8 and 9 were averaged across all ages and transmission rates, so it should also be considered that *AZ dose (n1)* would be expected to have more influence at the higher transmission rates. Analysis of sensitivity to findings can be conducted for specific scenarios, e.g., for a specific age group and under a specific transmission scenario.

**Table 3.** Sensitivity of outcome values to changes in inputs (shown as cases per million). Minimum and maximum represent the smallest and largest values for each outcome node when selecting different states of the input nodes.

|  |  |  | Input nodes |  |  |  |  |
| --- | --- | --- | --- | --- | --- | --- | --- |
|  |  |  | AZ dose (n1) | Age group (n2) | Variant (n3) | Community transmission rate at x% over 6 months (n4) | Sex (n5) |
| Output nodes | *Die from vaccine-induced TTS (n15)* | Range | 1.18 | 0.19 | - | - | - |
|  |  | Min | 0.00 | 0.31 | - | - | - |
|  |  | Max | 1.18 | 0.50 | - | - | - |
|  | *Die from background CVST (n16)* | Range | - | 0.03 | - | - | - |
|  |  | Min | - | 0.03 | - | - | - |
|  |  | Max | - | 0.05 | - | - | - |
|  | *Die from background PVT (n17)* | Range | - | 0.53 | - | - | - |
|  |  | Min | - | 0.00 | - | - | - |
|  |  | Max | - | 0.53 | - | - | - |
|  | *Die from Covid (n18)* | Range | 215.92 | 655.53 | 89.39 | 269.85 | 16.24 |
|  |  | Min | 7.99 | 0.00 | 80.63 | 0.00 | 76.98 |
|  |  | Max | 223.91 | 655.53 | 170.02 | 269.85 | 93.22 |
|  | *Die from Covid-related CVST (n19)* | Range | 0.08 | 0.09 | 0.02 | 0.28 | 0.05 |
|  |  | Min | 0.05 | 0.04 | 0.07 | 0.00 | 0.06 |
|  |  | Max | 0.13 | 0.13 | 0.09 | 0.28 | 0.11 |
|  | *Die from Covid-related PVT (n20)* | Range | 0.90 | 1.04 | 0.23 | 3.04 | 0.39 |
|  |  | Min | 0.55 | 0.44 | 0.74 | 0.00 | 0.76 |
|  |  | Max | 1.45 | 1.48 | 0.97 | 3.04 | 1.16 |

Based on Table 3, it is evident that *Age group (n1)* is the most influential node for the *Die from Covid (n18)* node, as indicated by the large range of probabilities associated with this input. In contrast *Community transmission at x% over 6 months (n4)* was more influential than *Age group (n1)* for the outcomes relating to dying from COVID-19-related atypical severe blood clots (n19 & n20).

The results of each outcome under the best-case and worst-case input scenarios (defined in Supplementary S8 – Best-case and worst-case scenarios) are given in Table 4. Note that for best-case scenarios, males were used instead females for the *Die from Covid (n18)* outcome due to zero deaths in females in the younger age groups and low, rather than no transmission has been used for COVID-19-related outcomes. Delta variant was used in all scenarios to reflect the current situation in Australia. The largest range in values was for the *Die from Covid (n18)* outcome, comparing the worst-case scenario of unvaccinated males age >70 years in a high transmission scenario (5391 deaths per million) to the best-case scenario of fully vaccinated females aged 10-19 in a low transmission scenario (0.008 deaths per million). The outcome nodes that were least sensitive to changes in the input nodes (in absolute numbers) related to the risk of dying from CVST, either background rates (n16) or COVID-related (n19), where the minimum and maximum values varied by less than 1 death per million (0.024 and 0.445 respectively).

**Table 4.** Values of outcomes nodes (in cases per million) under best-case and worst-case scenarios. Scenario definitions are provided in Supplementary S8.

|  | *Die from vaccine induced TTS (n15)* | *Die from background CVST (n16)* | *Die from PVT background (n17)* | *Die from Covid (n18)* | *Die from Covid related CVST (n19)* | *Die from Covid related PVT (n20)* |
| --- | --- | --- | --- | --- | --- | --- |
| Best case | 0.000 | 0.027 | 0.000 | 0.008 | 0.001 | 0.006 |
| Worst case | 1.350 | 0.051 | 0.529 | 5391.488 | 0.446 | 8.518 |
| Range | 1.350 | 0.024 | 0.529 | 5391.480 | 0.445 | 8.512 |

## 4 DISCUSSION

BNs provided an ideal framework for rapidly prototyping a decision-support tool to consolidate the existing evidence on risks and benefits of the AZ vaccine. The BN for AZ vaccine risk-benefit analysis developed in this study enables users to set scenarios by vaccination status (none, one or two doses of AZ vaccine), age, gender, SARS CoV-2 variant, and community transmission rate. Parameterisation of the model for the Australian population was based on seven separate data sources. By helping the subjects matter experts to combine the relevant evidence from different sources, the resulting model can be used to probabilistically estimate and compare risks of adverse outcomes and generate meaningful scenarios for a risk-benefit analysis. The analyses also provide useful information for informing the debate on the risks of vaccine-induced TTS relative to the risks of dying from COVID-19 or associated CVST or PVT.

Using a BN framework to collate and analyse existing evidence highlighted several key messages for informing the risk-benefit analysis for the AZ vaccine. The rate of community transmission was found to be a major moderating influence on the risk-benefit analysis for the AZ vaccine. These results should therefore be considered in the context of the dynamic nature of the COVID-19 pandemic, where transmission rates are likely to change rapidly.

As expected, the model indicated that age was found to have a higher influence than sex on the risk of dying from COVID-19. While there was large variance in the risk of dying from COVID-19 depending on a person’s age, sex and vaccination status, there was less variance in the risk of developing and dying from either the background or COVID-19-related CVST or PVT. Knowing which outcomes were highly variable, as well as which inputs were highly influential on each outcome provides useful information when looking to design a simple decision-support tool for risk-benefit analysis. As an example, these details could be used to customise the inputs required to simplify the tool for the user (e.g. not asking for sex if it isn’t relevant). The same information could also be used to target certain groups, such as focusing efforts on certain age groups, or by those who have only had one vaccine dose.

One key advantage of BNs as modelling tools is the ability to easily update the model with new evidence by editing the CPTs (Fenton et al. 2013). This feature has proven to be crucial for our study, where new evidence was available weekly and needed to be updated on a regular basis. The transparency of BN models and the model-building process was also critical to building a trusted model. First, having a model that users can interrogate and see why a probability has been estimated for a given scenario allows users to explore and understand the model. Fully documenting the design assumptions and explicitly identifying the sources used for populating the CPTs allows users to understand and evaluate where the probabilities have originated from. Another advantage of BNs was the ability to easily compartmentalise the model into sub-models, and narrow or broaden the scope as required by the addition or removal of nodes. This capability facilitated a prototyping approach that was able to reflect the evolving evidence around the AZ vaccine.

Although the probabilities in the model were derived from empirical studies or publicly available government data, decisions about which data to include (based on availability, robustness and compatibility with other datasets), and how they should be integrated were based on expert judgement. The experts’ role in the process was therefore crucial in both selecting and interpreting the available evidence, such as when choosing between results of different studies. In some cases, it may be more appropriate to include the study with the larger cohort, whereas in other instances, a smaller study that more closely resembles the population being modelled was preferable. In other instances, expert knowledge was required to align the data from different sources. For example, the youngest age group reported for the data on the chance of developing TTS after each vaccine dose was for those under 50 years of age, whereas the model has five ten-year age groups for this same population (0-9 years, 10-19 years, etc.).

An important distinction when designing a model for use in the context of public health is whether it should be interpreted as an individual model or a population model. For example, does 10% chance of infection represent a 10% chance that an individual will become infected, or that 10% of the population will become infected? While in practice these values might be considered to be interchangeable, in this model, the distinction was important in the definition of the node states and the corresponding calculation of the probability of developing TTS. The individual model was designed to represent the probability of TTS after the first and second dose as independent events. To model population-level estimates, e.g., the number of cases of TTS per million people where 35% have had only one dose, and 35% have had two doses, the model should be designed to consider cumulative risk, i.e., those who received two doses were also exposed to the risk associated with the first dose (Lau et al. 2021). It is also important to note that individual factors, such as comorbidities and access to health care, were not included in the model, so the risk for a particular individual may vary substantially from the population estimate. As with any statistical model, communicating a clear interpretation of the BN is crucial in a decision-support context where clinicians and the public, as well as policy makers, public health managers and the broader scientific community might misinterpret the model outputs.

Constructing an evidence-based model relying on expert-derived assumptions introduced several challenges for validating the model. Whereas the predictive performance of a data-driven model can be validated using cross-validation, and expert-derived models can be validated based on the opinion of independent experts (Marcot 2017), neither option was suitable for the model presented here. Instead, the logic in the model was validated against independent calculations based on the same evidence. While this did not validate the evidence or the assumptions, it did confirm that the model accurately reflects the information reported in the selected sources.

The modeller-led approach used in this research, although well-suited to a rapid design process, was a deviation from best-practice expert-elicitation processes (for example Richards et al. 2013; Wu et al. 2021), where the initial conceptual model would ideally be developed by the subject matter experts, facilitated by the modelling experts. Instead, presenting the subject-matter experts with a draft BN structure based on preliminary discussions allowed for an expedited design process. The design process was also facilitated by structured questionnaires used to help the experts collate the evidence into a suitable format that the modelling team could use to populate the CPTs.

A key benefit of BNs is that the interface facilitates model interaction, allowing users to explore different scenarios and develop a deep understanding of how concepts are represented and related in the model. However, a BN interface can be daunting to untrained users and interpreting the probabilities in relatable terms can be difficult. A frontend interface for the model has been created to simplify the inputs and communicate the results in terms of relatable risk (Immunisation Coalition 2021). This publicly available decision-support tool can also be updated as the scope of the model is expanded to include other vaccines, adverse events and comorbidities.

While the scope of the present study is currently limited to the AZ vaccine, the process described here can be easily repeated to expand the model to include additional inputs, including other COVID-19 variants or vaccines, demographic variables such as remoteness, or individual variables such as comorbidities. Specific next steps in model development are the inclusion of long COVID as an outcome and the chance of myocarditis from mRNA vaccines. More generally, the process could be extended to consideration of vaccine safety in the context of other health outcomes, particularly in rapidly changing environments.

Although the model has been quantified using data from Australia as well as international data from the UK and US, it has been adapted for the Australian context, and priority has been given to Australian data if available. Using the same process described here, however, the model is easily adaptable to international settings by re-parameterising it with local data where available. Through rapid aggregation, modelling and communication of vaccine-related risks, the comparative merits of vaccination in target populations can be better understood, leading to improved decision-making by policy makers and public health managers, and increased capability and capacity for clinicians to guide patients to make informed decisions about vaccines.

## Data Availability

All data produced in the present work are contained in the manuscript.

## 5 ACKNOWLEDGMENTS

We thank Kim Sampson from Immunisation Coalition for facilitating the collaboration between authors, and A/Prof Hassan Valley (La Trobe University, Melbourne, Australia) for contributions to discussions about risk communication and data visualisation. Our Bayesian network model was built using GeNIe Modeler (BayesFusion 2019), available free of charge for academic research and teaching use from https://www.bayesfusion.com, and we thank Marek Druzdel for software support, in particular with supplementary material S2.We thank the THANZ Vaccine-induced Immune Thrombotic Thrombocytopenia (VITT) advisory group (in particular: Dr Anoop K Enjeti, A/Prof Vivien Chen, Prof Huyen Tran, A/Prof Jennifer Curnow, Dr Ibrahim Tohidi and Dr Danny Hsu) for their feedback about TTS related data.

