## Supplementary for "Designing an evidence-based Bayesian network for estimating the risk versus benefits of AstraZeneca COVID-19 vaccine"

### Supplementary S1 – Example questionnaire for evidence gathering

| **Q1. Effectiveness of AstraZeneca vaccine** | | | | | |
| --- | --- | --- | --- | --- | --- |
| Questions 1.1 to 1.5 relate to the effectiveness of the **AstraZeneca vaccine**. Please answer the following questions in relation to people of **all age groups** (not just those eligible for vaccination in Australia), keeping in mind that the evidence may be derived from overseas studies. | | | | | |
|  | Thinking about overall effectiveness in **all ages**: | **a) COVID-19-related death** | **b) COVID-19-related symptomatic infection** | **c) Severe COVID-19** | **Source of evidence (e.g. publications, WHO, ATAGI)** |
| **1.1** | What is the effectiveness (in %) of **one dose of the AZ vaccine** against a) COVID-19-related death, b) COVID-19-related ICU admission, and c) Severe COVID-19? |  |  |  |  |
| **1.2** | What is the effectiveness (in %) of **two doses of the AZ vaccine** against a) COVID-19-related death, b) COVID-19 related ICU admission, and c) Severe COVID-19? |  |  |  |  |
| **1.3** | Is there evidence to suggest that vaccine effectiveness against the different outcomes (a to c) differ between **age groups**? (If yes, further age-related questions will be asked in the next round.) |  |  |  |  |
| **1.4** | Is there evidence to suggest that vaccine effectiveness against the different outcomes (a to c) differ by **sex**? (If yes, further sex-related questions will be asked in the next round.) |  |  |  |  |

### Supplementary S2 - Explanation of Euclidian Distance in BNs

Euclidean distance is a simple measure of distance inspired by Euclidean geometry that calculates the distance between two points in Euclidean space by taking the square root of squares of differences along each of the dimensions. There is a simple interpretation of Euclidean distance when calculating the distance between two discrete probability distributions. We treat each of the outcomes of a random variable as a dimension, calculate squares of differences in each of the probabilities of individual states between the two distributions and, finally, take the square root of the sum of these squares of differences to calculate the distance between the two distributions.

Euclidean distance can be used to calculate the strength of the influence of a parent node on a child node. Consider the following fictitious example relating whether or not a person is vaccinated to their chance of developing a mild or severe infection.

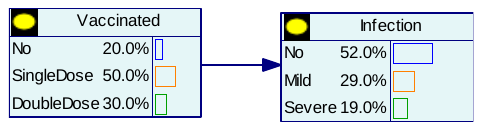

The conditional probability table for the *Infection* node is

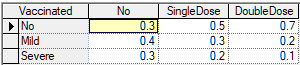

The influence of the node *Vaccinated* on *Infection* can be expressed as the difference between the conditional probability distributions over the child node under different values of the parent node.

In our example, there are three values of the parent node, *Vaccinated* = *No*, *Vaccinated* = *SingleDose*, and *Vaccinated* = *DoubleDose*. There are three sets of Euclidian distances for three sets of conditional probability distributions over the node *Infection* under these three values. Consider the distance between the cases *DoubleDose* and *No*. Squared differences between the probabilities of each of the states of the node *Infection* are as follows:

|  |  | Difference in probability based on parent node state | Squared difference |
| --- | --- | --- | --- |
| **Infection** | No | 0.7 – 0.3 = 0.4 | 0.16 |
|  | Mild | 0.2 – 0.4 = -0.2 | 0.04 |
|  | Severe | 0.1 - 0.3 = -0.2 | 0.04 |

The Euclidean distance between the two distributions is then Sqrt(0.16+0.04+0.04) = Sqrt(0.24) = 0.489897949. Similarly, we calculate the distance between the cases *DoubleDose* and *SingleDose* to be 0.244948974 and between *No* and *SingleDose* to be 0.244948974.

GeNIe normalizes these distances to the interval [0,1] by dividing each of them by the square root of 2, which is the largest possible value of an unnormalised Euclidean distance between two probability distributions. Normalised distances are in our example: 0.346410162, 0.173205081, and 0.173205081.

Now, the strength of the influence of *Vaccination* on *Infection* can be expressed by the average distance (0.346410162 + 0.173205081 + 0.173205081)/3 =0.230940108, and by the maximum distance Max(0.346410162, 0.173205081, 0.173205081) = 0.346410162.

### Supplementary S3 – Values for conditional probability tables

| **AZ dose (n1)** | |
| --- | --- |
| None | 0.3 |
| One | 0.35 |
| Two | 0.35 |

| **Age group (n2)** | |
| --- | --- |
| Age 0-9 | 0.12387612 |
| Age 10-19 | 0.11988012 |
| Age 20-29 | 0.14085914 |
| Age 30-39 | 0.14585415 |
| Age 40-49 | 0.12787213 |
| Age 50-59 | 0.12187812 |
| Age 60-69 | 0.1048951 |
| Age 70+ | 0.11488511 |

| **SARS-CoV-2 variant (n3)** | |
| --- | --- |
| Alpha/ancestral | 0.05 |
| Delta | 0.95 |

| **Community transmission at x% over 6 months (n4)** | |
| --- | --- |
| None (0) | 0.1 |
| ATAGI Low | 0.1 |
| ATAGI Medium | 0.1 |
| ATAGI High | 0.1 |
| One % | 0.1 |
| Two % | 0.1 |
| NSW 200 daily | 0.1 |
| NSW 1000 daily | 0.1 |
| VIC 1000 daily | 0.1 |
| QLD 1000 daily | 0.1 |

| **Sex (n5)** | |
| --- | --- |
| Male | 0.5 |
| Female | 0.5 |

| **Vaccine-induced TTS (n6)** | | | | | | | | |
| --- | --- | --- | --- | --- | --- | --- | --- | --- |
| AZ doses received | None | | | | | | | |
| Age | 0-9 | 10-19 | 20-29 | 30-39 | 40-49 | 50-59 | 60-69 | 70+ |
| Yes | 0 | 0 | 0 | 0 | 0 | 0 | 0 | 0 |
| No | 1 | 1 | 1 | 1 | 1 | 1 | 1 | 1 |
| AZ doses received | One | | | | | | | |
| Age | 0-9 | 10-19 | 20-29 | 30-39 | 40-49 | 50-59 | 60-69 | 70+ |
| Yes | 2.50E-05 | 2.50E-05 | 2.50E-05 | 2.50E-05 | 2.50E-05 | 2.70E-05 | 1.60E-05 | 1.85E-05 |
| No | 0.999975 | 0.999975 | 0.999975 | 0.999975 | 0.999975 | 0.999973 | 0.999984 | 0.9999815 |
| AZ doses received | Two | | | | | | | |
| Age | 0-9 | 10-19 | 20-29 | 30-39 | 40-49 | 50-59 | 60-69 | 70+ |
| Yes | 1.80E-06 | 1.80E-06 | 1.80E-06 | 1.80E-06 | 1.80E-06 | 1.80E-06 | 1.80E-06 | 1.80E-06 |
| No | 0.9999982 | 0.9999982 | 0.9999982 | 0.9999982 | 0.9999982 | 0.9999982 | 0.9999982 | 0.9999982 |

| **CVST over 6 weeks (n7)** | | | | | | | | |
| --- | --- | --- | --- | --- | --- | --- | --- | --- |
| Age | 0-9 | 10-19 | 20-29 | 30-39 | 40-49 | 50-59 | 60-69 | 70+ |
| Yes | 3.80E-07 | 3.80E-07 | 6.40E-07 | 6.40E-07 | 6.40E-07 | 7.50E-07 | 7.50E-07 | 7.30E-07 |
| No | 0.99999962 | 0.99999962 | 0.99999936 | 0.99999936 | 0.99999936 | 0.99999925 | 0.99999925 | 0.99999927 |

| **PVT over 6 weeks (n8)** | | | | | | | | |
| --- | --- | --- | --- | --- | --- | --- | --- | --- |
| Age | 0-9 | 10-19 | 20-29 | 30-39 | 40-49 | 50-59 | 60-69 | 70+ |
| Yes | 0.00E+00 | 0.00E+00 | 2.00E-07 | 2.60E-07 | 5.60E-07 | 9.10E-07 | 1.76E-06 | 1.96E-06 |
| No | 1 | 1 | 0.9999998 | 0.99999974 | 0.99999944 | 0.99999909 | 0.99999824 | 0.99999804 |

| **Vaccine effectiveness against symptomatic infection (n9)** | | | | | | |
| --- | --- | --- | --- | --- | --- | --- |
| Variant | Alpha/ancestral | | | Delta | | |
| AZ doses | None | One | Two | None | One | Two |
| Effective | 0 | 0.6 | 0.8 | 0 | 0.33 | 0.61 |
| Ineffective | 1 | 0.4 | 0.2 | 1 | 0.67 | 0.39 |

| **Vaccine effectiveness against death if infected (n10)** | | | | | | |
| --- | --- | --- | --- | --- | --- | --- |
| Variant | Alpha/ancestral | | | Delta | | |
| AZ doses | None | One | Two | None | One | Two |
| Effective | 0 | 0.8 | 0.95 | 0 | 0.69 | 0.9 |
| Ineffective | 1 | 0.2 | 0.05 | 1 | 0.31 | 0.1 |

| **Risk of symptomatic infection by age and variant (n11)** | | | | | | | | | | | | | | | |  |  |  |  |
| --- | --- | --- | --- | --- | --- | --- | --- | --- | --- | --- | --- | --- | --- | --- | --- | --- | --- | --- | --- |
| Variant | Alpha/ancestral | | | | | | | | | | | | | | |  |  |  |  |
| Age | 0-9 | 10-19 | | 20-29 | | 30-39 | | 40-49 | | 50-59 | | 60-69 | | 70+ | |  |  |  |  |
| Yes | 0.0416611 | 0.0702759 | | 0.1589055 | | 0.1212599 | | 0.1005435 | | 0.0977633 | | 0.0812794 | | 0.1126984 | |  |  |  |  |
| No | 0.9583389 | 0.9297241 | | 0.8410945 | | 0.8787401 | | 0.8994565 | | 0.9022367 | | 0.9187206 | | 0.8873016 | |  |  |  |  |
| Variant | Delta | | | | | | | | | | | | | | |  |  |  |  |
| Age | 0-9 | 10-19 | | 20-29 | | 30-39 | | 40-49 | | 50-59 | | 60-69 | | 70+ | |  |  |  |  |
| Yes | 0.09182884 | 0.1423347 | | 0.15400685 | | 0.11311761 | | 0.09119044 | | 0.09033166 | | 0.05484361 | | 0.04305606 | |  |  |  |  |
| No | 0.90817116 | 0.8576653 | | 0.84599315 | | 0.88688239 | | 0.90880956 | | 0.90966834 | | 0.94515639 | | 0.95694394 | |  |  |  |  |
| **Risk of symptomatic infection under current transmission and vaccination status (n12)** | | | | | | | | | | | | | | | | | | | |
| Risk of symptomatic infection by age and variant | | | Yes | | | | | | | | | | | | | | | | |
| Vaccine effectiveness against symptomatic infection | | | Effective | | | | | | | | | | | | | | | | |
| Community transmission at x% over 6 months | | | None (0) | | ATAGI High | | ATAGI Medium | | ATAGI Low | | 1 % | | 2 % | | NSW 200 daily | | NSW 1000 daily | VIC 1000 daily | QLD 1000 daily |
| Yes | | | 0 | | 0 | | 0 | | 0 | | 0 | | 0 | | 0 | | 0 | 0 | 0 |
| No | | | 1 | | 1 | | 1 | | 1 | | 1 | | 1 | | 1 | | 1 | 1 | 1 |
| Risk of symptomatic infection by age and variant | | | Yes | | | | | | | | | | | | | | | | |
| Vaccine effectiveness against symptomatic infection | | | Ineffective | | | | | | | | | | | | | | | | |
| Community transmission at x% over 6 months | | | None (0) | | ATAGI High | | ATAGI Medium | | ATAGI Low | | 1 % | | 2 % | | NSW 200 daily | | NSW 1000 daily | VIC 1000 daily | QLD 1000 daily |
| Yes | | | 0 | | 0.00471 | | 0.04469 | | 0.5759 | | 0.1 | | 0.2 | | 0.04455 | | 0.22276 | 0.27289 | 0.35067 |
| No | | | 1 | | 0.99529 | | 0.95531 | | 0.4241 | | 0.9 | | 0.8 | | 0.95545 | | 0.77724 | 0.72711 | 0.64933 |
| Risk of symptomatic infection by age and variant | | | No | | | | | | | | | | | | | | | | |
| Vaccine effectiveness against symptomatic infection | | | Effective | | | | | | | | | | | | | | | | |
| Community transmission at x% over 6 months | | | None (0) | | ATAGI High | | ATAGI Medium | | ATAGI Low | | 1 % | | 2 % | | NSW 200 daily | | NSW 1000 daily | VIC 1000 daily | QLD 1000 daily |
| Yes | | | 0 | | 0 | | 0 | | 0 | | 0 | | 0 | | 0 | | 0 | 0 | 0 |
| No | | | 1 | | 1 | | 1 | | 1 | | 1 | | 1 | | 1 | | 1 | 1 | 1 |
| Risk of symptomatic infection by age and variant | | | No | | | | | | | | | | | | | | | | |
| Vaccine effectiveness against symptomatic infection | | | Ineffective | | | | | | | | | | | | | | | | |
| Community transmission at x% over 6 months | | | None (0) | | ATAGI High | | ATAGI Medium | | ATAGI Low | | 1 % | | 2 % | | NSW 200 daily | | NSW 1000 daily | VIC 1000 daily | QLD 1000 daily |
| Yes | | | 0 | | 0 | | 0 | | 0 | | 0 | | 0 | | 0 | | 0 | 0 | 0 |
| No | | | 1 | | 1 | | 1 | | 1 | | 1 | | 1 | | 1 | | 1 | 1 | 1 |

| **CVST from COVID-19 (n13)** | | | | |
| --- | --- | --- | --- | --- |
| Risk of symptomatic infection under current transmission and vaccination status | Yes | | No | |
| Sex | Male | Female | Male | Female |
| Yes | 2.89E-05 | 5.42E-05 | 0 | 0 |
| No | 0.99997113 | 0.9999458 | 1 | 1 |

| **PVT from COVID-19 (n14)** | | | | |
| --- | --- | --- | --- | --- |
| Risk of symptomatic infection under current transmission and vaccination status | Yes | | No | |
| Sex | Male | Female | Male | Female |
| Yes | 4.83E-04 | 3.18E-04 | 0 | 0 |
| No | 0.9995174 | 0.99968159 | 1 | 1 |

| **Die from vaccine-induced TTS (n15)** | | |
| --- | --- | --- |
| TTS 31-08 | Yes | No |
| Yes | 0.05 | 0 |
| No | 0.95 | 1 |

| **Die from CVST (n16)** | | |
| --- | --- | --- |
| CVST over 6 weeks | Yes | No |
| Yes | 0.07 | 0 |
| No | 0.93 | 1 |

| **Die from PVT (n17)** | | |
| --- | --- | --- |
| PVT over 6 weeks | Yes | No |
| Yes | 0.27 | 0 |
| No | 0.73 | 1 |

| **Die from COVID-19 (n18)** | | | | | | | | |
| --- | --- | --- | --- | --- | --- | --- | --- | --- |
| Risk of symptomatic infection under current transmission and vaccination status | Yes | | | | | | | |
| Vaccine effectiveness against death if infected | Effective | | | | | | | |
| Sex | Male | | | | | | | |
| Age | 0-9 | 10-19 | 20-29 | 30-39 | 40-49 | 50-59 | 60-69 | 70+ |
| Yes | 0 | 0 | 0 | 0 | 0 | 0 | 0 | 0 |
| No | 1 | 1 | 1 | 1 | 1 | 1 | 1 | 1 |
| Risk of symptomatic infection under current transmission and vaccination status | Yes | | | | | | | |
| Vaccine effectiveness against death if infected | Effective | | | | | | | |
| Sex | Female | | | | | | | |
| Age | 0-9 | 10-19 | 20-29 | 30-39 | 40-49 | 50-59 | 60-69 | 70+ |
| Yes | 0 | 0 | 0 | 0 | 0 | 0 | 0 | 0 |
| No | 1 | 1 | 1 | 1 | 1 | 1 | 1 | 1 |
| Risk of symptomatic infection under current transmission and vaccination status | Yes | | | | | | | |
| Vaccine effectiveness against death if infected | Ineffective | | | | | | | |
| Sex | Male | | | | | | | |
| Age | 0-9 | 10-19 | 20-29 | 30-39 | 40-49 | 50-59 | 60-69 | 70+ |
| Yes | 0 | 0.00030321 | 0.00032457 | 0.00080402 | 0.00086505 | 0.00374787 | 0.01879699 | 0.21743389 |
| No | 1 | 0.99969679 | 0.99967543 | 0.99919598 | 0.99913495 | 0.99625213 | 0.98120301 | 0.78256611 |
| Risk of symptomatic infection under current transmission and vaccination status | Yes | | | | | | | |
| Vaccine effectiveness against death if infected | Ineffective | | | | | | | |
| Sex | Female | | | | | | | |
| Age | 0-9 | 10-19 | 20-29 | 30-39 | 40-49 | 50-59 | 60-69 | 70+ |
| Yes | 0 | 0 | 0 | 0.00044336 | 0.00062933 | 0.00285919 | 0.00806916 | 0.1910828 |
| No | 1 | 1 | 1 | 0.99955664 | 0.99937067 | 0.99714081 | 0.99193084 | 0.8089172 |
| Risk of symptomatic infection under current transmission and vaccination status | No | | | | | | | |
| Vaccine effectiveness against death if infected | Effective | | | | | | | |
| Sex | Male | | | | | | | |
| Age | 0-9 | 10-19 | 20-29 | 30-39 | 40-49 | 50-59 | 60-69 | 70+ |
| Yes | 0 | 0 | 0 | 0 | 0 | 0 | 0 | 0 |
| No | 1 | 1 | 1 | 1 | 1 | 1 | 1 | 1 |
| Risk of symptomatic infection under current transmission and vaccination status | No | | | | | | | |
| Vaccine effectiveness against death if infected | Effective | | | | | | | |
| Sex | Female | | | | | | | |
| Age | 0-9 | 10-19 | 20-29 | 30-39 | 40-49 | 50-59 | 60-69 | 70+ |
| Yes | 0 | 0 | 0 | 0 | 0 | 0 | 0 | 0 |
| No | 1 | 1 | 1 | 1 | 1 | 1 | 1 | 1 |
| Risk of symptomatic infection under current transmission and vaccination status | No | | | | | | | |
| Vaccine effectiveness against death if infected | Ineffective | | | | | | | |
| Sex | Male | | | | | | | |
| Age | 0-9 | 10-19 | 20-29 | 30-39 | 40-49 | 50-59 | 60-69 | 70+ |
| Yes | 0 | 0 | 0 | 0 | 0 | 0 | 0 | 0 |
| No | 1 | 1 | 1 | 1 | 1 | 1 | 1 | 1 |
| Risk of symptomatic infection under current transmission and vaccination status | No | | | | | | | |
| Vaccine effectiveness against death if infected | Ineffective | | | | | | | |
| Sex | Female | | | | | | | |
| Age | 0-9 | 10-19 | 20-29 | 30-39 | 40-49 | 50-59 | 60-69 | 70+ |
| Yes | 0 | 0 | 0 | 0 | 0 | 0 | 0 | 0 |
| No | 1 | 1 | 1 | 1 | 1 | 1 | 1 | 1 |

| **Die from COVID-19-related CVST (n19)** | | |
| --- | --- | --- |
| CVST from Covid | Yes | No |
| Yes | 0.174 | 0 |
| No | 0.826 | 1 |

| **Die from COVID-19-related PVT (n20)** | | |
| --- | --- | --- |
| PVT from Covid | Yes | No |
| Yes | 0.199 | 0 |
| No | 0.801 | 1 |

### Supplementary S4 – Probabilities for scenario

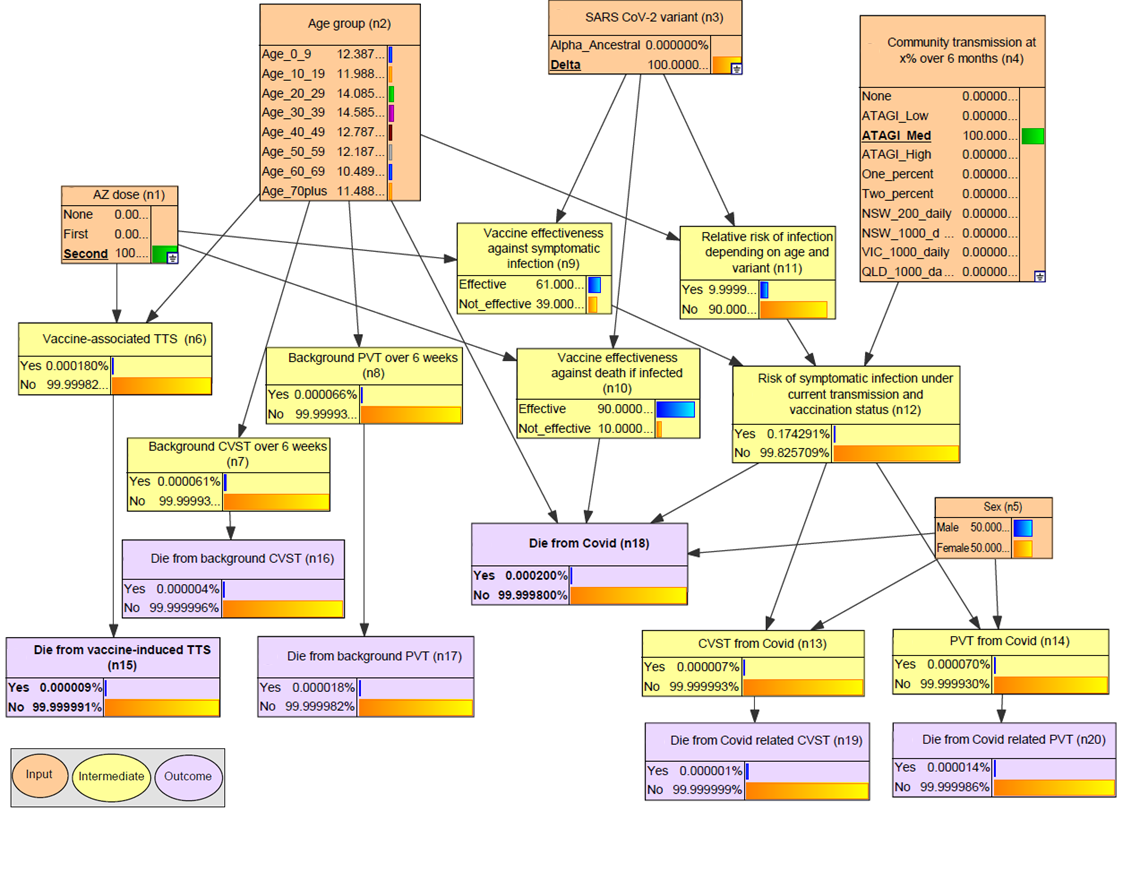

***Figure S4.1.*** *Probability values for a medium transmission scenario (n4) for the Delta variant (n3) and a fully vaccinated population (n1).*

### Supplementary S5 - Assumptions

Taken from: Lau, C. L., H. J. Mayfield, J. E. Sinclair, S. J. Brown, M. Waller, A. K. Enjeti, A. Baird, K. Short, K. Mengersen and J. Litt (2021). "Risk-benefit analysis of the AstraZeneca COVID-19 vaccine in Australia using a Bayesian network modelling framework." medRxiv: 2021.2009.2030.21264337.

**Table S4.1. Age distribution of reported cases of COVID-19 in NSW, 29/6/2021 to 13/8/2021.**

| **Age group** | **Cases** |
| --- | --- |
| 0-9* | 734 |
| 10-19* | 1100 |
| 20-29 | 1399 |
| 30-39 | 1064 |
| 40-49 | 752 |
| 50-59 | 710 |
| 60-69 | 371 |
| ≥70 | 319 |
| **Total** | **6449** |

*NSW Health reported 1834 cases in the 0-19 age group. Numbers based on assumption that 40% cases in the 0-19 age group were aged 0-9, and 60% were aged 10-19 age.

Sources:

NSW Government *NSW COVID-19 cases data*. 2021. <https://data.nsw.gov.au/nsw-covid-19-data/cases>.

Australian Government Department of Health *Cases and deaths by age and sex*. Coronavirus (COVID-19) case numbers and statistics, 2021. <https://www.health.gov.au/news/health-alerts/novel-coronavirus-2019-ncov-health-alert/coronavirus-covid-19-case-numbers-and-statistics#cases-and-deaths-by-age-and-sex>.

**Table S4.2. Age distribution of reported cases of COVID-19 in Australia, January to December 2020.**

| **Age group** | **Cases** |
| --- | --- |
| 0-9 | 1480 |
| 10-19 | 2416 |
| 20-29 | 6419 |
| 30-39 | 5072 |
| 40-49 | 3687 |
| 50-59 | 3417 |
| 60-69 | 2445 |
| ≥70 | 3713 |
| **Total** | **28649** |

Sources:

Australian Government Department of Health *COVID-19 summary statistics*. Coronavirus (COVID-19) case numbers and statistics, 2021. <https://www.health.gov.au/news/health-alerts/novel-coronavirus-2019-ncov-health-alert/coronavirus-covid-19-case-numbers-and-statistics#covid19-summary-statistics>.

Australian Government Department of Health *National Notifiable Diseases Surveillance System public datasets*. 2021. <https://www1.health.gov.au/internet/main/publishing.nsf/Content/ohp-pub-datasets.htm>.

**Table S4.3. Cases, deaths, and case fatality rate of COVID-19 in Australia by age and gender, January 2020 to 31/8/2021.**

|  | **Male** | | | **Female** | | |
| --- | --- | --- | --- | --- | --- | --- |
| **Age group** | **Cases** | **Deaths** | **Case fatality** | **Cases** | **Deaths** | **Case fatality** |
| 0-9 | 2848 | 0 | 0% | 2317 | 0 | 0% |
| 10-19 | 3298 | 1 | 0.03% | 3196 | 0 | 0% |
| 20-29 | 6162 | 2 | 0.03% | 5859 | 0 | 0% |
| 30-39 | 4975 | 4 | 0.08% | 4511 | 2 | 0.04% |
| 40-49 | 3468 | 3 | 0.09% | 3178 | 2 | 0.06% |
| 50-59 | 2935 | 11 | 0.37% | 2798 | 8 | 0.29% |
| 60-69 | 1862 | 35 | 1.88% | 1735 | 14 | 0.81% |
| ≥70 | 2042 | 444 | 21.74% | 2512 | 480 | 19.11% |
| **Total** | **27,226** | **500** | **1.84%** | **26,106** | **506** | **1.94%** |

Sources:

Australian Government Department of Health *COVID-19 summary statistics*. Coronavirus (COVID-19) case numbers and statistics, 2021. <https://www.health.gov.au/news/health-alerts/novel-coronavirus-2019-ncov-health-alert/coronavirus-covid-19-case-numbers-and-statistics#covid19-summary-statistics>.

Australian Government Department of Health *National Notifiable Diseases Surveillance System public datasets*. 2021. <https://www1.health.gov.au/internet/main/publishing.nsf/Content/ohp-pub-datasets.htm>.

**Table S4.4. Probability of infection (over 6 months) based on different intensities of community transmission.**

| **Intensity of community transmission** | **Cases per 100,000 over 16 weeks*** | **Cases per million over 6 months** | **Estimated % of population infected over 6 months** |
| --- | --- | --- | --- |
| Zero | 0 | 0 | 0% |
| Low* | 29 | 471 | 0.047% |
| Medium* | 275 | 4,469 | 0.447% |
| High* | 3,544 | 57,590 | 5.759% |
| 1% over 6 months |  | 10,000 | 1.0% |
| 2% over 6 months |  | 20,000 | 2.0% |
| 200 cases/day in NSW^a^ |  | 4,455 | 0.446% |
| 1000 cases/day in NSW^a^ |  | 22,276 | 2.228% |
| 1000 cases/day in VIC^b^ |  | 27,289 | 2.729% |
| 1000 cases/day in QLD^c^ |  | 35,067 | 3.507% |

***** Definitions of low, medium, and high transmission (cases per 100,000 over 16 weeks) as defined by ATAGI document ‘Weighing up the potential benefits and risk of harm from COVID-19 Vaccine AstraZeneca’. Low – similar to first wave in Australia. Medium – similar to second wave in VIC. High – similar to Europe in January 2021.

^a^Based on NSW population of 8.17 million. ^b^Based on VIC population of 6.67 million. ^c^Based on QLD population of 5.19 million.

Source:

Australian Government Department of Health *COVID-19 vaccination – Weighing up the potential benefits against risk of harm from COVID-19 Vaccine AstraZenec*. 2021. <https://www.health.gov.au/resources/publications/covid-19-vaccination-weighing-up-the-potential-benefits-against-risk-of-harm-from-covid-19-vaccine-astrazeneca>.

**Table S4.5. Relative risk of infection by age group and variant (chance of infection if overall probability of infection of 1% in all ages).**

| **Age group** | **Alpha/wild** | **Delta** |
| --- | --- | --- |
| 0-9 | 0.42% | 0.92% |
| 10-19 | 0.70% | 1.42% |
| 20-29 | 1.59% | 1.54% |
| 30-39 | 1.21% | 1.13% |
| 40-49 | 1.01% | 0.91% |
| 50-59 | 0.98% | 0.90% |
| 60-69 | 0.81% | 0.55% |
| ≥70 | 1.13% | 0.43% |
| **Overall** | **1.00%** | **1.00%** |

Sources:

Australian Government Department of Health COVID-19 summary statistics. Coronavirus (COVID-19) case numbers and statistics, 2021. https://www.health.gov.au/news/health- alerts/novel-coronavirus-2019-ncov-health-alert/coronavirus-covid-19-case-numbers-and- statistics#covid19-summary-statistics.

NSW Government NSW COVID-19 cases data. 2021. https://data.nsw.gov.au/nsw-covid-19- data/cases.

Australian Government Department of Health Cases and deaths by age and sex. Coronavirus (COVID-19) case numbers and statistics, 2021. https://www.health.gov.au/news/health- alerts/novel-coronavirus-2019-ncov-health-alert/coronavirus-covid-19-case-numbers-and- statistics#cases-and-deaths-by-age-and-sex.

Australian Government Department of Health National Notifiable Diseases Surveillance System public datasets. 2021. https://www1.health.gov.au/internet/main/publishing.nsf/Content/ohp-pub-datasets.htm.

**Table S4.6. Age distribution of Australian population, December 2020.**

| **Age group** | **Population** | **% of total population** |
| --- | --- | --- |
| 0-9 | 3,189,750 | 12.4% |
| 10-19 | 3,086,855 | 12.0% |
| 20-29 | 3,627,055 | 14.1% |
| 30-39 | 3,755,673 | 14.6% |
| 40-49 | 3,292,645 | 12.8% |
| 50-59 | 3,138,303 | 12.2% |
| 60-69 | 2,700,998 | 10.5% |
| ≥70 | 2,958,236 | 11.5% |
| **Total** | **25,749,515** | **100%** |

Source:

Australian Bureau of Statistics *Data downloads - time series spreadsheets*. National, state and territory population, 2021. <https://www.abs.gov.au/statistics/people/population/national-state-and-territory-population/dec-2020#data-download>.

**Table S4.7. Estimated background incidence and fatality of atypical blood clots (CVST and PVT combined) over 6 weeks (in populations who have not received AZ vaccine and not diagnosed with COVID-19).**

| **Age group** | **Incidence of atypical blood clots (CVST and PVT) over 6 weeks  (per million)** | **Fatality rates from atypical blood clots (CVST and PVT) over 6 weeks (per million)** | **Overall case fatality rate from atypical blood clots (CVST and PVT)** |
| --- | --- | --- | --- |
| 0-9 | 0.38 | 0.03 | 7.0% |
| 10-19 | 0.38 | 0.03 | 7.0% |
| 20-29 | 0.84 | 0.10 | 11.8% |
| 30-39 | 0.90 | 0.12 | 12.8% |
| 40-49 | 1.20 | 0.20 | 16.3% |
| 50-59 | 1.66 | 0.30 | 18.0% |
| 60-69 | 2.51 | 0.53 | 21.0% |
| ≥70 | 2.69 | 0.58 | 21.6% |

Source:

Espen Saxhaug Kristoffersen, et al., Incidence and Mortality of Cerebral Venous Thrombosis in a Norwegian Population. Stroke, 2020. 51(10): p. 3023-3029

W. Ageno, et al., Incidence rates and case fatality rates of portal vein thrombosis and Budd- Chiari Syndrome. Thromb Haemost, 2017. 117(4): p. 794-800

Søgaard, K. K., B. Darvalics, E. Horváth–Puhó and H. T. Sørensen (2016). "Survival after splanchnic vein thrombosis: A 20-year nationwide cohort study." Thrombosis Research 141: 1-7.

### Supplementary S6 - Manual calculations for validation

**Table S5.1. Results of manual calculations used to validate the mathematical assumptions used to parameterise the model. Values provided are KM (blue), MW (green) and model estimates (purple).**

| **Assumptions:** Variant: Delta  Vaccine coverage: 70% has first dose, 35% had two doses | | | |
| --- | --- | --- | --- |
| Questions | | Calculated estimate | |
| **1.** For a 30-39 year old male, what is the chance of symptomatic infection under ATAGI high transmission if: | a) Not vaccinated | 0.065144400  0.06514443 | |
|  | b) Had one dose | 0.043646800  0.043550000  0.043646769 | |
|  | c) Had two doses | 0.025406300  0.025350000  0.025406328 | |
| **2.** For a 30-39 year old male, what is the overall chance of dying from COVID-19 under ATAGI high transmission if: | a) Not vaccinated | 0.000052377  0.000052000  0.000052377 | |
|  | b) Had one dose | 0.000010879  0.000010912  0.000010879 | |
|  | c) Had two doses | 0.000002043  0.000002000  0.000002043 | |
| *** 3.** For one million 50-59 year old females, if 70% had one dose, and 35% had two doses: | a) How many cases of vaccine-induced TTS would we expect? | 19.530000000  19.5300000000  19.53 | |
|  | b) How many TTS-related deaths would we expect? | 0.976500000  0.9765000000  0.977 | |
| **4.** For one million 70+ year old males, if 70% had one dose, and 35% had two doses: | a) How many symptomatic cases would we expect over 6 months if there was ATAGI medium transmission during this time? | 1291.1200  1289.7291  1291.1216 | |
|  | b) How many deaths would we expect over 6 months if there was ATAGI medium transmission during this time? | 161.63900  161.43988  161.63938 | |
| **5.** If a 60-69 year old female was diagnosed with COVID-19: |  | CVST | PVT |
|  | a) What are her chances of developing Covid related atypical blood clots (CVST and PVT)? | 0.000054197  0.0000542000  0.0000541969 | 0.000318407  0.0003180000  0.000318407 |
|  | b) What are her chances of dying from Covid related atypical blood clots (CVST and PVT)? | 0.000009430  0.0000094308  0.0000094302 | 0.000063363  0.0000632820  0.0000633629 |
| Additional population risk: | c) What are her background chances of developing atypical blood clots (CVST and PVT)? | 0.000000750  0.00000075 | 0.000001760  0.00000176 |
|  | d) What are her background chances of dying from atypical blood clots (CVST and PVT)? | 0.000000053  0.0000000525 | 0.000000475  0.0000004752 |

*Model estimates for Question 3 are calculated using the population level model described in section 3.2

### Supplementary S7 - Influence of inputs nodes on outcomes

**Table S6.1 Sensitivity analysis of effect of changes to outcomes nodes based on states of input nodes (shown as cases per million).**

|  |  | **Die from vaccine-associated TSS (n15)** | **Die from background CVST risk (n16)** | **Die from background PVT (n17)** | **Die from Covid (n18)** | **Die from Covid related CVST (n19)** | **Die from Covid related PVT (n20)** |
| --- | --- | --- | --- | --- | --- | --- | --- |
| Average |  | 0.444 | 0.043 | 0.178 | 85.100 | 0.087 | 0.087 |
| AZ dose (n1) | None | 0.000 | 0.043 | 0.178 | 223.910 | 0.131 | 1.447 |
|  | One | 1.178 | 0.043 | 0.178 | 43.231 | 0.086 | 0.950 |
|  | Two | 0.090 | 0.043 | 0.178 | 7.989 | 0.050 | 0.551 |
| Age group (n2) | 0-9 | 0.469 | 0.027 | 0.000 | 0.000 | 0.078 | 0.863 |
|  | 10-19 | 0.469 | 0.027 | 0.000 | 1.470 | 0.121 | 1.339 |
|  | 20-29 | 0.469 | 0.045 | 0.054 | 1.744 | 0.134 | 1.480 |
|  | 30-39 | 0.469 | 0.045 | 0.070 | 4.930 | 0.099 | 1.088 |
|  | 40-49 | 0.469 | 0.045 | 0.151 | 4.768 | 0.080 | 0.879 |
|  | 50-59 | 0.504 | 0.053 | 0.246 | 20.864 | 0.079 | 0.870 |
|  | 60-69 | 0.312 | 0.053 | 0.475 | 52.395 | 0.049 | 0.536 |
|  | 70+ | 0.355 | 0.051 | 0.529 | 655.528 | 0.040 | 0.439 |
| SARS-CoV-2 variant (n3) | Alpha / ancestral | 0.444 | 0.043 | 0.178 | 170.025 | 0.067 | 0.737 |
|  | Delta | 0.444 | 0.043 | 0.178 | 80.630 | 0.088 | 0.971 |
| Community transmission at x% over 6 months (n4) | None | 0.444 | 0.043 | 0.178 | 0.000 | 0.000 | 0.000 |
|  | Low | 0.444 | 0.043 | 0.178 | 2.207 | 0.002 | 0.025 |
|  | Med | 0.444 | 0.043 | 0.178 | 20.940 | 0.021 | 0.236 |
|  | High | 0.444 | 0.043 | 0.178 | 269.849 | 0.276 | 3.043 |
|  | 1% | 0.444 | 0.043 | 0.178 | 46.857 | 0.048 | 0.528 |
|  | 2% | 0.444 | 0.043 | 0.178 | 93.714 | 0.096 | 1.057 |
|  | 200 NSW | 0.444 | 0.043 | 0.178 | 20.875 | 0.021 | 0.235 |
|  | 1000 NSW | 0.444 | 0.043 | 0.178 | 104.378 | 0.107 | 1.177 |
|  | 2000 NSW | 0.000 | 0.000 | 0.000 | 0.000 | 0.000 | 0.000 |
|  | 1000 VIC | 0.444 | 0.043 | 0.178 | 127.868 | 0.131 | 1.442 |
|  | 1000 QLD | 0.444 | 0.043 | 0.178 | 164.313 | 0.168 | 1.853 |

### Supplementary S8 – Best-case and worst-case scenarios

**Table S7.1: Best-case and worst-case scenarios for each outcome node in terms of probability of dying. Note that males have been used instead of females for the *‘Die from Covid’* outcome due to zero deaths in females in the younger age groups. Low, rather than no transmission has been used for COVID-19-related outcomes.**

|  | Best case | Worst case |
| --- | --- | --- |
| Die from vaccine induced TTS (n15) | Age group = 50-59 AZ dose = None | Age group = 50-59  AZ dose = First |
| Die from Background CVST (n16) | Age group = 0-19 | Age group = 70+ |
| Die from background PVT (n17) | Age group = 0-19 | Age group = 70+ |
| Die from Covid (n18) | Transmission = ATAGI Low,  Age group = 10-19,  AZ dose = Second,  Variant = Delta,  Sex = Male | Transmission = ATAGI High,  Age group = 70+,  AZ dose = None,  Variant = Delta,  Sex = Male |
| Die from Covid-related CVST (n19) | Transmission = ATAGI Low,  Age group = 60-69,  AZ dose = Second,  Variant = Delta,  Sex = Female | Transmission = ATAGI High,  Age group = 20-29,  AZ dose = Second,  Variant = Delta,  Sex = Male |
| Die from Covid-related PVT (n20) | Transmission = ATAGI Low,  Age group = 60-69,  AZ dose = Second,  Variant = Delta,  Sex = Female | Transmission = ATAGI High,  Age group = 20-29,  AZ dose = Second,  Variant = Delta,  Sex = Male |
